# Infection fatality rate of SARS-CoV-2 infection in a German community with a super-spreading event

**DOI:** 10.1101/2020.05.04.20090076

**Authors:** Hendrik Streeck, Bianca Schulte, Beate M. Kümmerer, Enrico Richter, Tobias Höller, Christine Fuhrmann, Eva Bartok, Ramona Dolscheid, Moritz Berger, Lukas Wessendorf, Monika Eschbach-Bludau, Angelika Kellings, Astrid Schwaiger, Martin Coenen, Per Hoffmann, Birgit Stoffel-Wagner, Markus M. Nöthen, Anna-Maria Eis-Hübinger, Martin Exner, Ricarda Maria Schmithausen, Matthias Schmid, Gunther Hartmann

**Affiliations:** Institute of Virology, University Hospital, University of Bonn, Germany, and German Center for Infection Research (DZIF), partner site Bonn-Cologne; Institute for Hygiene and Public Health, University Hospital, University of Bonn, Germany; Institute for Medical Biometry, Informatics and Epidemiology, University Hospital, University of Bonn, Germany; Institute of Clinical Chemistry and Clinical Pharmacology, University Hospital, University of Bonn, Germany; German Center for Infection Research (DZIF), partner site Bonn-Cologne; Clinical Study Core Unit, Study Center Bonn (SZB), Institute of Clinical Chemistry and Clinical Pharmacology, University Hospital, University of Bonn, Germany; Biobank Core Unit, University Hospital, University of Bonn, Germany; Institute of Human Genetics, University Hospital, University of Bonn, Germany

## Abstract

The world faces an unprecedented SARS-CoV2 pandemic where many critical factors still remain unknown. The case fatality rates (CFR) reported in the context of the SARS-CoV-2 pandemic substantially differ between countries. For SARS-CoV-2 infection with its broad clinical spectrum from asymptomatic to severe disease courses, the infection fatality rate (IFR) is the more reliable parameter to predict the consequences of the pandemic. Here we combined virus RT-PCR testing and assessment for SARS-CoV2 antibodies to determine the total number of individuals with SARS-CoV-2 infections in a given population.

**Methods:** A sero-epidemiological GCP- and GEP-compliant study was performed in a small German town which was exposed to a super-spreading event (carnival festivities) followed by strict social distancing measures causing a transient wave of infections. Questionnaire-based information and biomaterials were collected from a random, household-based study population within a seven-day period, six weeks after the outbreak. The number of present and past infections was determined by integrating results from anti-SARS-CoV-2 IgG analyses in blood, PCR testing for viral RNA in pharyngeal swabs and reported previous positive PCR tests.

**Results:** Of the 919 individuals with evaluable infection status (out of 1,007; 405 households) 15.5% (95% CI: [12.3%; 19.0%]) were infected. This is 5-fold higher than the number of officially reported cases for this community (3.1%). Infection was associated with characteristic symptoms such as loss of smell and taste. 22.2% of all infected individuals were asymptomatic. With the seven SARS-CoV-2-associated reported deaths the estimated IFR was 0.36% [0.29%; 0.45%]. Age and sex were not found to be associated with the infection rate. Participation in carnival festivities increased both the infection rate (21.3% vs. 9.5%, p<0.001) and the number of symptoms in the infected (estimated relative mean increase 1.6, p=0.007). The risk of a person being infected was not found to be associated with the number of study participants in the household this person lived in. The secondary infection risk for study participants living in the same household increased from 15.5% to 43.6%, to 35.5% and to 18.3% for households with two, three or four people respectively (p<0.001).

**Conclusions:** While the number of infections in this high prevalence community is not representative for other parts of the world, the IFR calculated on the basis of the infection rate in this community can be utilized to estimate the percentage of infected based on the number of reported fatalities in other places with similar population characteristics. Whether the specific circumstances of a super-spreading event not only have an impact on the infection rate and number of symptoms but also on the IFR requires further investigation. The unexpectedly low secondary infection risk among persons living in the same household has important implications for measures installed to contain the SARS-CoV-2 virus pandemic.

## Introduction

The novel SARS-CoV-2 coronavirus causing a respiratory disease termed COVID-19^1,2^ is affecting almost every country worldwide^3^. One of the reasons for its rapid spread is its ability to transmit already during the asymptomatic phase of infection, reported to be responsible for approximately 40% of SARS-CoV-2 transmission events^4,5^. As the COVID-19 pandemic continues to grow in extent, severity, and socio-economic consequences, its fatality rate remains unclear. SARS-CoV-2 infection presents with a broad spectrum of clinical courses from asymptomatic to fatal, complicating the definition of a ‘case’. About 80-91% of the infections have been reported to show only mild to moderate symptoms including sore throat, dry cough and fever^6^. These are currently often left undiagnosed. Together with different PCR test capacities and different regulations for testing, the ratio of SARS-CoV-2-associated deaths to overall reported cases (case fatality rate, CFR) inherently differs between countries ^7^. The current estimate of the CFR in Germany by the World Health Organization (WHO) is between 2.2% and 3.4%^3^. The data basis, however, for calculating the CFR is weak, with the consequence that epidemiological modeling is currently associated with a large degree of uncertainty. However, epidemiological modeling is urgently needed to design the most appropriate prevention and control strategies to counter the pandemic and to minimize collateral damage to societies.

Unlike the CFR, the infection fatality rate (IFR) includes the whole spectrum of infected individuals, from asymptomatic to severe. The IFR is recommended as a more reliable parameter than the CFR for evidence-based assessment of the SARS-CoV-2 pandemic (Center for Evidence-Based Medicine, CEBM in Oxford). The IFR includes infections based on both PCR testing and virus-specific antibodies. Mild and moderate disease courses are also included, which tend to not be captured and documented by PCR testing alone. Active infections before seroconversion are included into IFR-calculation by PCR testing. In this testing scheme, only those individuals may be missed who already became negative in the PCR test but have not yet reached antibody levels above the threshold of the antibody detection assay^8^.

Recently commercial assays became available with a specificity of up to 99% to allow for a reliable serological analysis of SARS-CoV-2-specific antibodies^9^. Of note, lower specificities of ELISA tests reported in the literature are partially due to the use of beta versions of the ELISA and to different calculation algorithms (>0.3 log(Ig ratio)) defining positive values^10^. Furthermore, even an assay with a validated specificity of 99% has limitations in its accuracy to reliably identify infected individuals in populations with low seroprevalence (e.g. <1 %). We chose the community of Gangelt, where due to a super-spreading event, the officially reported cases were 3% (time of study period). In this community, carnival festivities around February 15^th^ were followed by a massive outbreak of SARS-CoV-2 infections. Strict measures including a suggested curfew were immediately taken to slow down further spreading of the infection. Given its relatively closed community with little tourism and travel, this community was identified as an ideal model to better understand SARS-CoV2 spreading, prevalence of symptoms, as well as the infection fatality rate. The results presented here were obtained in the context of the larger study program termed COVID-19-Case-Cluster Study. The parts of a larger study program which are presented here were specifically designed to determine the total number of infected and the IFR. In addition, the spectrum of symptoms, as well as the associations with age, sex, household size, co-morbidities and participation in carnival festivities, were examined.

## Materials and methods

### Study design

This study was conducted between March 31^st^, 2020 and April 6^th^, 2020 in Gangelt, a community with 12,597 inhabitants (as of Jan 1^st^, 2020) located in the German county of Heinsberg in North Rhine-Westphalia. For this cross-sectional epidemiological study, all inhabitants of Gangelt were eligible. Enrollment was based on a sample of 600 persons contained in the Heinsberg civil register (“Melderegister”), which is the public authority that collects all names and addresses of the inhabitants of Gangelt. All study participants provided written and informed consent before enrolment. For children under 18 years, written and informed consent was provided by the persons with care and custody of the children following aged-adapted participant information. In addition to the data provided by the study participants, aggregated data on mortality and socio-demographic characteristics were collected. The latter data were provided by the district administration of Heinsberg and the Statistics & IT Service of the German federal state of North Rhine-Westphalia. The study was approved by the Ethics Committee of the Medical Faculty of the University of Bonn (approval number 085/20) and has been registered at the German Clinical Trials Register (https://www.drks.de, identification number DRKS00021306). The study was conducted in accordance with good clinical (GCP) and epidemiological practice (GEP) standards and the Declaration of Helsinki.

### Sampling and procedures

Based on the sample size recommendations of the World Health Organisation (WHO) (see below), the aim was to collect data from at least 300 households in Gangelt. To reach this target, a sample of 600 persons aged older than 18 years was drawn from the civil register. Sampling was done randomly under the side condition that all 600 persons had different surnames, as it was assumed that different surnames were likely to indicate different households. After sampling, the 600 selected persons were contacted by mail and were invited to the study acquisition center, which was established at the site of a public school in Gangelt. The letters sent to the 600 selected persons also included invitations for all persons living in the respective households to participate in the study. Persons aged older than 80 years or immobile were offered the opportunity to be visited at home. After having provided written and informed consent, study participants completed a questionnaire querying information including demographics, symptoms, underlying diseases, medication and participation in carnival festivities (main carnival session “Kappensitzung” and others). Furthermore, study participants were asked to provide blood specimens and pharyngeal swabs. Blood was centrifuged and EDTA-plasma was stored until analysis (−80°C). Analyses were performed in batches at the central laboratory of the University Hospital Bonn (UKB), which is accredited according to DIN EN ISO 15189:2014. Anti-SARS-CoV-2 IgA and Anti-SARS-CoV-2 IgG were determined with enzyme-linked immunosorbent assays (ELISA) on the EUROIMMUN Analyzer I platform (most recent CE version for IgG ELISA as of April 2020, specificity 99.1%, sensitivity 90.9%, data sheet as of April 7, 2020, validated in cooperation with the Institute of Virology of the Charité in Berlin, and the Erasmus MC in Rotterdam, Euroimmun, Lübeck, Germany). The data sheet (April 7, 2020) reports cross-reactivities with anti-SARS-CoV-1-IgG-antibodies, but not with MERS-CoV-, HCoV-229E-, HCoV-NL63-, HCoV-HKU1- or HCoV-OC43-IgG antibodies. In our study, infected included positives (ratio of 1.1 or higher, 91% positive in neutralization assay) and equivocal positives (ratio 0.8 to 1.1, 56% positive in neutralization assays). Assays were performed in line with the guidelines of the German Medical Association (RiliBÄK) with stipulated internal and external quality controls. Pharyngeal swabs were stored in UTM Viral Stabilization Media at 4 °C at the study acquisition center for up to four hours. The cold chain remained uninterrupted during transport. At the Institute of Virology of the UKB swab samples were homogenized by short vortexing, and 300 μl of the media containing sample were transferred to a sterile 1.5 ml microcentrifuge tube and stored at 4 °C. Viral RNA was extracted on the chemagic™ Prime™ instrument platform (Perkin Elmer) using the chemagic Viral 300 assay according to manufacturer’s instructions. The RNA was used as template for three real time RT-PCR reactions (SuperScript™III One-Step RT-PCR System with Platinum™ TaqDNA Polymerase, Thermo Fisher) to amplify sequences of the SARS-CoV-2 E gene (primers E_Sarbeco_F1 and R, and probe E_Sarbeco_P1^11^), the RdRP gene (primers RdRP_SARSr_F, and R, and probe RdRP_SARSr-P2^11^), and an internal control for RNA extraction, reverse transcription, and amplification (innuDETECT Internal Control RNA Assay, Analytik Jena #845-ID-0007100). Samples were considered positive for SARS-CoV-2 if amplification occurred in both virus-specific reactions. All PCR protocols and materials were used according to clinical diagnostics standards and guidelines of the Virology Diagnostics Department of the UKB. Neutralization assays were performed using a SARS-CoV-2 strain isolated in Bonn from a throat swab of a patient from Heinsberg. Plasma samples from study participants were inactivated at 56°C for 30 min. In a first round, neutralizing activity was analyzed by a microneutralization test using 100 TCID_50_ similar as described^12^. Serial 2-fold dilutions (starting dilution 1:2, 50 μl per well) of plasma were performed and mixed with equal volumes of virus solution. All dilutions were made in DMEM (Gibco) supplemented with 3% fetal bovine serum (FBS, Gibco) and each plasma dilution was run in triplicate. After incubation for 1 h at 37°C, 2×10^4^ Vero E6 cells were added to each well and the plates were incubated at 37°C for 2 days in 5% CO2 before evaluating the cytopathic effect (CPE) via microscopy. In each experiment, plasma from a SARS-CoV-2 IgG negative person was included and back titration of the virus dilution was performed. Titers were calculated according to the Spearman-Kaerber formula^13^ and are presented as the reciprocals of the highest plasma dilution protecting 50% of the wells. To further assess the neutralizing activity of plasma samples exhibiting neutralizing antibody titers below 2.8 in the microneutralization test, a plaque reduction neutralization test was performed. To this end, heat inactivated plasma samples were serially two-fold diluted starting with 1: 2 up to 1:1,024. 120 μl of each plasma dilution was mixed with 100 plaque forming units (PFU) of SARS-CoV-2 in 120 μl OptiPRO^TM^SFM (Gibco) cell culture medium. After incubation of 1 h at 37°C, 200 μl of each mixture were added to wells of a 24 well plate seated the day before with 1.5×10^5^ Vero E6 cells/well. After incubation for 1 h at 37°C, the inoculum was removed and cells were overlayed with a 1:1 mixture of 1.5% carboxymethylcellulose (Sigma) in 2×MEM (Biochrom) with 4% FBS (Gibco). After incubation at 37°C for three days in 5% CO2, the overlay was removed and the 24 well plates were fixed using a 6% formaldehyde solution and stained with 1% crystal violet in 20% ethanol.

### Data management and quality control

Planning and conduct of the study were supported by the Clinical Study Core Unit (Studienzentrale) of the Study Centre Bonn (SZB). Support included protocol and informed consent development following specifications of the World Health Organization with regards to pandemic events^14^, data management, submission to the ethics committee, clinical trial monitoring and quality control. Study data were collected and managed using REDCap electronic data capture tools hosted at Institute for Medical Biometry, Informatics and Epidemiology^13, 14^. REDCap (Research Electronic Data Capture) is a secure, web-based software platform designed to support data capture for research studies, providing 1) an intuitive interface for validated data capture; 2) audit trails for tracking data manipulation and export procedures; 3) automated export procedures for seamless data downloads to common statistical packages; and 4) procedures for data integration and interoperability with external sources. Questionnaire data were recorded on site using paper case report forms and were entered into the electronic study database using double data entry by trained study personnel. Comparisons between entries were made by the data management unit of the SZB; non-matches were corrected, and duplicated entries were deleted, after assessing the original paper case report forms. Additionally, plausibility checks of demographic data were performed. Study personnel were trained with respect to informed consent and study procedures prior to inclusion of first study participant. The study team was supported on site in Gangelt by a quality control manager who refined workflow processes and monitored critical processes such as obtaining informed consent. Furthermore, regulatory advice could be given whenever asked for or needed. Data entry personnel was trained for double data entry prior to data entry and only then granted database access authorization. Contact with the responsible data managers could be established when needed. Diagnostic data were imported into the trial database automatically via validated interfaces. Following the completion of the study, critical data was monitored by an experienced clinical trial monitor which included (but was not limited to) a check of availability of source data (completed questionnaires), random source data verification of diagnostic data and a check of signatures of all informed consent forms obtained.

### Statistical analysis

In the absence of any pilot data on SARS-CoV-2 infection rates in Gangelt, sample size calculations were based on the WHO population-based age-stratified seroepidemiological investigation protocol for COVID-19 virus infection^14^. According to the recommendations stated in the protocol, a size of 200 samples is sufficient to estimate SARS-CoV-2-prevalence rates <10% with an expected margin of error (defined by the expected width of the 95% confidence interval associated with the seroprevalence point estimate obtained using binomial likelihood) smaller than 10%. In order to rule out larger margins of error due to dependencies of persons living in the same household and to be able to analyze seroprevalence (i.e., infection rates) also in subgroups defined by participant age, it was planned to recruit 1,000 participants living in at least 300 households. Statistical analysis was carried out by two independently working statisticians (MS, MB) using version 3.6.1 of the R Language for Statistical Computing (R Core Team 2019: R: A Language and Environment for Statistical Computing. R Foundation for Statistical Computing, Vienna, Austria) and version 9.4 of the SAS System for Windows (copyright © 2002-2012 by SAS Institute Inc., Cary, NC, USA). Participants with a missing anti-SARS-CoV-2 IgG/A or PCR test result were excluded from analysis, as they were not evaluable for infection status (Fig. 1B). Participants that did not report a previous positive PCR test result were documented as PCR_rep_ negative. Missing and unknown values in the co-morbidity and symptom variables were not imputed, as listwise deletion reduced sample sizes by less than 5%. Age groups were formed according to the classification system of the *Robert Koch Institute* (RKI), which is the German federal government agency and research institute responsible for infectious disease control and prevention.

**Fig. 1.**
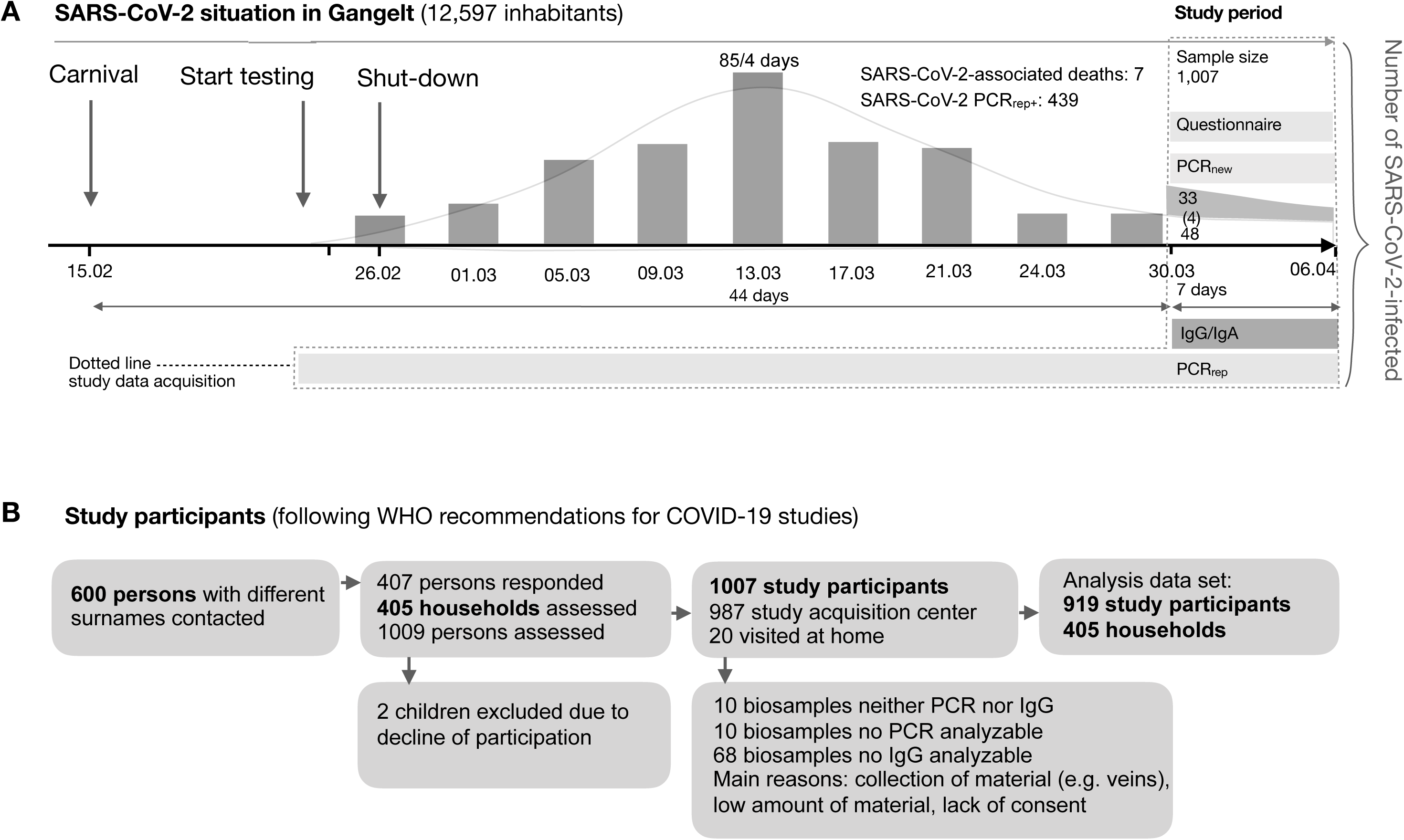
**A:** Timeline of the SARS-CoV-2 superspreading event. On February 15^th^, 2020, a carnival celebration became a SARS-CoV2 superspreading event in the German community of Gangelt. The resulting increase in SARS-CoV-2-infected people was instantly countered by a complete shutdown (schools, restaurants, stores, etc.). As a result, the number of reported cases (PCR_rep_) reached its peak around March 13 (85 reported in a four-day period) and declined thereafter, with 48 new cases reported in the seven-day study period (March 30 to April 6). Thus, at the starting point of the study (March 30), the main wave of new infections had already passed. The number of PCR-positive cases found in the study population (PCR_new_) was 33 (four of those reported PCR positive in the past). This situation in the community of Gangelt was ideal to assess the cumulative real number of SARS-CoV-2-infected individuals (area within dotted line: PCR_rep_, PCR_new_ and anti-SARS-CoV-2 IgG/A). **B:** *Enrolment and flow of participants through the study*.

Descriptive analyses included the calculation of means (plus standard deviations, sds) and medians (plus minimum and maximum values) for continuous variables, and numbers (n, with percentages) for categorical variables. Associations between continuous variables were analyzed using the Pearson correlation coefficient (r).

Generalized estimation equations (GEE)^15^ with exchangeable correlation structure within household clusters were used to adjust point estimates and confidence intervals (CIs) for possible dependencies between participants living in the same household. By definition, GEE models employ quasi-likelihood methods to obtain point estimates and CIs. Adjustments for possible sex and age effects were made by including these variables as additional covariables in the GEE models. One person of diverse sex (Table 1) was excluded from the models including sex as covariable. For binary outcomes (e.g. infection status), GEE models with a logistic link function were applied. Results of logistic GEE models are presented in terms of either back-transformed mean estimates (GEE models with a single covariable) or odds ratios (ORs, GEE models with ≥ 1 covariables). For count data (e.g. number of symptoms), Poisson GEE models with a logarithmic link function were used. Results of Poisson GEE models are presented in terms of either back-transformed mean estimates (GEE models with a single covariable) or estimated relative mean increases/decreases (GEE models with ≥ 1 covariables). For all GEE models, the estimated correlation between participants living in the same household cluster (rho) is reported. On a household-level basis (with households assumed to be independent sampling units), quasi-Poisson models with offset values defined by the logarithmized household cluster size were applied. Wald tests were used to test covariables for statistical significance.

**Table 1.** Characteristics and co-morbidities of the 919 study participants in 405 households evaluable for infection status.

| Characteristic | Value |
| --- | --- |
| Size of household clusters – no. (%) |  |
| 1 pers | 98 (24.20) |
| 2 pers | 184 (45.43) |
| 3 pers | 59 (14.57) |
| 4 pers | 49 (12.10) |
| 5 pers or more | 15 (3.70) |
| Age |  |
| Median (range) | 53 yr (1 yr – 90 yr) |
| Distribution – no. (%) |  |
| <5 yr | 6 (0.65) |
| 5-14 yr | 55 (5.98) |
| 15-34 yr | 176 (19.15) |
| 35-59 yr | 344 (37.43) |
| 60-79 yr | 266 (28.94) |
| >79 yr | 72 (7.83) |
| Sex – no. (%) |  |
| Male | 451 (49.08) |
| Female | 467 (50.82) |
| Diverse | 1 (0.11) |
| IgG – no. (%) |  |
| High | 106 (11.53) |
| Intermediate | 19 (2.07) |
| Normal | 794 (86.40) |
| PCR<sub>new</sub> – no. (%) |  |
| Positive | 33 (3.59) |
| Negative | 886 (96.41) |
| PCR<sub>reported</sub> – no. (%) |  |
| Yes | 117 (12.79) |
| No | 796 (86.99) |
| Not known | 2 (0.22) |
| Missings: 4 |  |
| PCR<sub>reported</sub> positive – no (%) |  |
| Yes | 22 (18.97) |
| No | 93 (80.17) |
| Not known | 1 (0.86) |
| Missings: 1 |  |
| Lung disease – no (%) |  |
| Yes | 107 (1.67) |
| No | 798 (87.02) |
| Not known | 12 (1.31) |
| Missings: 2 |  |
| Cardiovascular disease – no (%) |  |
| Yes | 128 (13.93) |
| No | 779 (84.77) |
| Not known | 12 (1.31) |
| Missings: 0 |  |
| Neurological disease - no (%) |  |
| Yes | 41 (4.46) |
| No | 875 (95.21) |
| Not known | 3 (0.33) |
| Missings: 0 |  |
| Cancer – no(%) |  |
| Yes | 69 (7.52) |
| No | 842 (92.82) |
| Not known | 6 (0.65) |
| Missings: 2 |  |
| Diabetes – no (%) |  |
| Yes | 79 (8.62) |
| No | 832 (90.73) |
| Not known | 6 (0.65) |
| Missings: 2 |  |
| Carnival – no (%) |  |
| Yes | 417 (45.52) |
| No | 498 (54.37) |
| Not known | 1 (0.11) |
| Missings: 3 |  |

All CIs presented in this work were computed using the 95% level. CIs are Wald CIs and were not adjusted for multiple comparisons unless otherwise stated. All statistical hypothesis tests were two-sided; p-values < 0.05 were considered significant. The Bonferroni-Holm procedure was applied to adjust p-values for multiple comparisons as indicated.

Infection rates obtained from IgG and IgA measurements were additionally corrected for possible misclassification bias using the matrix method^16^, with sensitivity and specificity values obtained from the ELISA manufacturer’s (Euroimmun, Lübeck, Germany) validation data sheet (version: April 7, 2020). No adjustments were made for age and sex, as these variables were not found to be associated with infection status (Fig. 6A). To account for possible clustering effects due to participants living in the same household, confidence intervals for the corrected infection rate estimates were computed using a cluster bootstrap procedure with 10,000 bootstrap samples^17^. With this procedure, household clusters were sampled with replacement. Within sampled clusters, no additional resampling of household members was carried out. The distributions of the bootstrapped corrected infection rate estimates were symmetrical and close to normality (as indicated by normal quantile-quantile plots), and the percentile method was applied to calculate CI limits. Note: Throughout the paper, the term *rate* refers to the number of persons experiencing an event divided by the number of the reference population, in line with the definition of the IFR^18^. We adopted this definition due to its widespread use in the context of COVID-19 research, keeping in mind that “rates” are usually defined in terms of person-time (e.g. Rothman et al^19^).

## Results

### Study design and study population

The major objective of this study was to determine the total number of individuals infected by SARS-CoV-2 in the given defined population. This number together with the reported SARS-CoV-2-associated fatalities in that same population allows the calculation of the infection fatality rate (IFR, according to Centre for Evidence-Based Medicine, CEBM, Oxford University, to be distinguished from case fatality rate, CFR). In the German community Gangelt (12,597 inhabitants), a super-spreading event (carnival festivities incl. “Kappensitzung” on February 15, 2020), was followed by numerous measures starting February 28 (shut-down) to limit the further spread of infections (**Fig. 1A**). This local infection hotspot was closely monitored by health authorities, and a high PCR test rate revealed an increase in officially reported cases, with a maximum around March 13 when 85 individuals tested PCR positive for SARS-CoV-2 in a 4-day period. Numbers declined afterwards down to 48 PCR positive cases officially reported during the 7-day period of the present study (March 30^th^ - April 6^th^), not counting the 33 new PCR positives detected in this study. The total number of officially reported PCR positives on April 6^th^ was 388, also excluding the 33 PCR positives of this study. By the end of the 7-day study period, a total of 7 SARS-CoV-2-positive individuals had died in the community of Gangelt since the super-spreading event (average age 80.8 years, sd ± 3.5 years). In January, February and March 2020, a total of 48 people died in Gangelt, which was 3 people more than in the same period the year before. At the start date of data and material acquisition of the study, 340 PCR positives were reported in the community which is 2.7% of the population.

Our study design addressed the recommendations for COVID-19 studies by the WHO^14^. For the study, 600 adult persons with different surnames in Gangelt were randomly selected, and all household members were asked to participate in the study. Data and materials were collected over a 7-day period (March 30 to April 6) six weeks after super-spreading event. Of the 1,007 individuals participating in the study, 987 individuals were seen in the local study acquisition center in a community school, and 20 individuals were visited in their homes due to age or limited mobility. Complete information from both pharyngeal swabs and blood samples was available for 919 study participants living in 405 households **(Fig. 1B)**. The demographic characteristics of the study participants, including age, sex and the number of participants living in the same household, are summarized in **Table 1**. The comparison of age groups in the study population to the community Gangelt, to the state North Rhine-Westphalia, (NRW) and to Germany is illustrated in **supplementary figure 1**. Characteristics of the 88 study participants who were not evaluable for infection status, mainly children due to lack of biomaterials, are provided in the **supplementary table**.

### Number of SARS-CoV-2-infected and infection fatality rate (IFR)

The analysis of IgA and IgG levels measured in plasma samples of all study participants by ELISA (Euroimmun) showed a positive correlation (r=0.778, CI 95%: [0.751-0.802]: **Fig. 2A**). While 18.50% of all study participants were found to be IgA positive, 13.60% were IgG positive (**Fig. 2B**). Correction for sensitivity and specificity of the ELISA (specificity 99.1%, sensitivity 90.9%) revealed a much lower corrected value of 10.63% [7.48%; 13.88%] for IgA and a slightly higher value of 14.11% [11.15%; 17.27%] for IgG (**Fig. 2B**). The higher specificity of the IgG ELISA (99.1%, validation reported April 7, 2020 by company based on 1,656 samples) compared to IgA ELISA (91.2%) was confirmed by our own independent analysis of control samples (specificity 98.3%: 1 positive in 68 samples of healthy control individuals, 1 positive in 32 samples of patients with cardiovascular disease, 0 positive in 9 samples of 7 patients with PCR-confirmed infection with endemic coronaviruses). To illustrate the difference between a specificity of 99% and 98%, the correction for specificity of 98% is added in light gray (**Fig. 2B,C**). Based on these data, a “seropositive” study participant was defined as being positive for IgG (mean of values corrected for sensitivity and specificity of all study participants; **Fig. 2B**). The neutralization activity of IgG-positive plasma samples was analyzed using a microneutralization assay combined with a plaque reduction neutralization test. Results are shown in **Suppl. Fig. 2**.

**Fig. 2.**
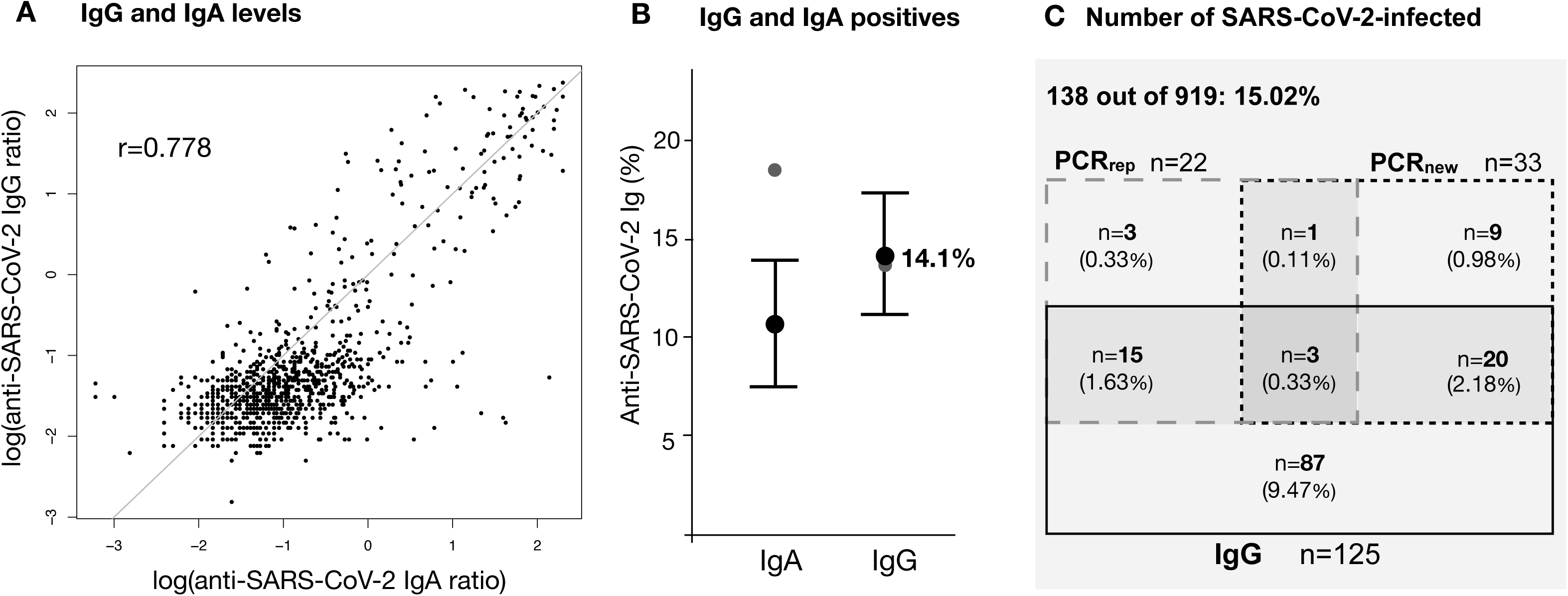
IgA and IgG levels and number of SARS-CoV2 infected in the study population. IgA and IgG were quantified (Euroimmun ELISA) in single plasma samples obtained from the study participants at one time point during the seven-day acquisition period of the study. **A:** IgG plotted against IgA in plasma of 919 study participants (log-scale, r = 0.778). The gray line indicates equality of log(IgA) and log(IgG). **B:** Estimated percentage of IgA reactives (> 0.8; black circle: 10.63% [7.48%; 13.88%]; gray circle: raw sample proportion 170/919, 18.50%) and IgG reactives (>0.8; black circle: 14.11% [11.15% - 17.27%]; gray circle: raw sample proportion, 125/919, 13.60%). Estimates were corrected for household clustering (cluster bootstrap) and for sensitivity and specificity (matrix method) of the IgA (sensitivity 100%; specificity 91.2%) and IgG (sensitivity 90.9%; specificity 99.1%) ELISA (validation sheet version of April 7, 2020). **C:** The absolute numbers of IgG reactives (rectangle with black border), PCR_new_ positives (rectangle with dashed border, left side) and PCR_rep_ positives (rectangle with dashed border, right side) as well as the respective overlaps of values are depicted (percentages in brackets). The number of infected (total grey area) is defined as study participants positive for at least one of either IgG, PCR_new_ or PCR_rep_ (138/919, 15.02%; raw percentages not corrected for sensitivity and specificity of the ELISA).

To determine the total number of infected individuals, all study participants were tested for the presence of virus in pharyngeal swabs by SARS-CoV-2 PCR in addition to serology. Of the 919 participants of the study, 33 tested positive (PCR_new_: 3.59%). Furthermore, based on the information collected from the questionnaire, 22 study participants reported that they had had a SARS-CoV-2 positive PCR test in the past (PCR_rep_: 2.39%). The combination of serology (non-corrected IgG values) and past and present PCR testing yielded a total number of 138 study participants (15.02%) that had been previously or were at that time point infected by SARS-CoV-2 as illustrated in **Fig. 2C**. The inclusion of IgG values corrected for sensitivity and specificity in the calculation resulted in an estimated 15.53% [12.31%; 18.96%] cumulative SARS-CoV-2-infected of all study participants.

To determine the infection fatality rate (IFR), the estimated infection rate of 15.53% in the study population was applied to the total population in the community (12,597) yielding an estimated number of 1,956 [1,551; 2,389] infected people. With 7 SARS-CoV-2-associated deaths, as reported to the authors by the local administration, the estimated IFR was 7/1,956 = 0.00358 [0.00293; 0.00451] (0.358% [0.293%; 0.451%]) (**Fig. 3A**) at the end of the acquisition period. While the percentage of previously reported cases as collected from the questionnaire in the study population was 2.39% (PCR_rep+_), the percentage of officially reported cases in the community of Gangelt at the end of the study period (April 6) was 3.08% (388/12,597). This indicates that previously SARS-CoV-2 diagnosed individuals were somewhat underrepresented in our study, possibly due to previously diagnosed people not opting to participate in the study given their known infection status, or for other reasons, such as quarantine, not feeling well or hospitalization. Thus, applying the corresponding correction factor (3.08% / 2.39% = 1.29) to the infection rate of 15.53% of our study population, the resulting corrected infection rate was 19.98% [15.84%; 24.40%] (**Fig. 3B**). Accordingly, the corrected higher infection rate reduced the IFR to an estimated 0.278% [0.228%; 0.351 %] (**Fig. 3C**).

**Fig. 3.**
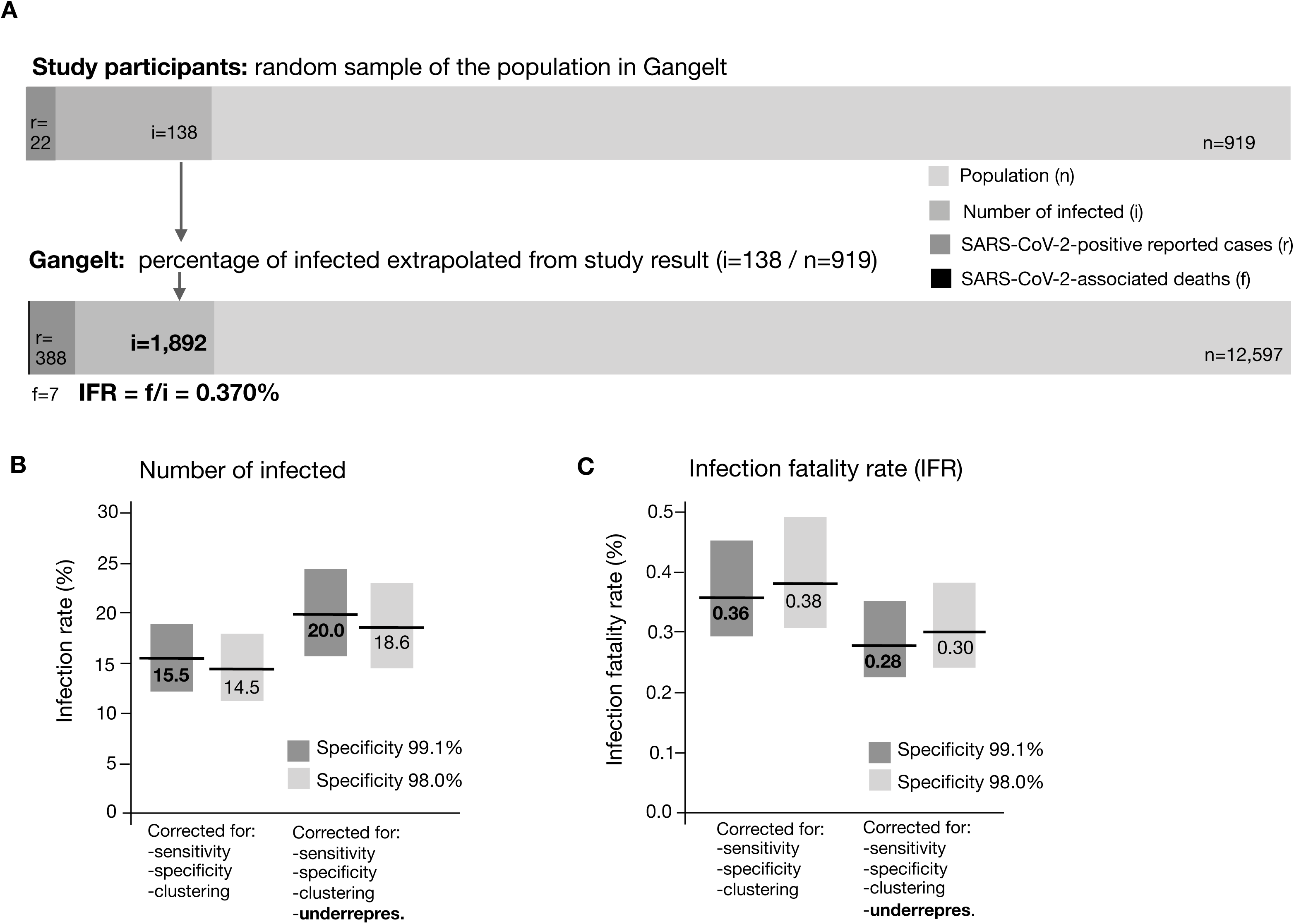
Estimation of the SARS-CoV2 infection rate and the IFR. **A:** The number of SARS-CoV-2-positive reported cases in the study population is known from the questionnaire (r = 22). The observed number of infected in the study population is known from the data available (at least one of either IgG+, PCR_new_+ or PCR_rep_+, i = 138). The ratio of i / n (study participants, 919) = 0.1502 is a raw estimate of the number of infected in the whole population of Gangelt (i = 0.1502 × 12,597 ≈ 1,892). A raw estimate of the IFR in Gangelt is therefore given by the number of SARS-CoV-2-associated deaths (f = 7) / (i = 1,892) = 0.370%. **B:** The infection rate estimated from the IgG and PCR data in the study population, corrected for both sensitivity/specificity of the ELISA (matrix method) and household clustering (cluster bootstrap), is 15.53% [12.31%; 18.96%] (left bar, dark gray). An additional correction was made for the underrepresentation of reported PCR positive (PCR_rep_+) in the study population (22/919 = 0.0239) as compared to the real proportion of PCR_rep_+ in Gangelt (388/12,597 = 0.0308), increasing the infection rate by the factor 0.0308/0.0239 = 1.2866 to 19.98% [15.84%; 24.40%] (third bar from left, dark gray). The bars in light gray depict the values corrected for a theoretical specificity of the ELISA of 98% (light gray) instead of the 99% provided on the data sheet of the company. **C:** Infection fatality rate calculated based on the estimated infection rates and the number of SARS-CoV-2-associated deaths (7 by the end of the acquisition period, mean age 81.1 ± 3.3 years, age range 78 years to 85 years). Similar to the infection rates in B, the estimated IFR of 0.36% [0.29%; 0.45%] (left bar) may be an estimate at the upper limit of the real IFR in Gangelt. IFR estimates were obtained by dividing the number of SARS-CoV-2-associated deaths (7) by the point estimates and 95% CI limits of the infection rates in B.

### Infection rate, symptoms and intensity of disease

A number of symptoms have been reported to be associated with SARS-CoV-2 infection^1^. In the questionnaire, study participants were asked to indicate whether they experienced any of the described symptoms since the beginning of the pandemic February 15^th^. Noting that symptoms may vary in both frequency (**Table 2**) and intensity, and that causal relationships cannot be established by a cross-sectional study, the following symptoms were found to be significantly associated with SARS-CoV-2 infection (based on IgG^+^, PCR_rep+_, PCR_new+_, ranked by odds ratios with 95% CIs, adjusted for sex and age, Bonferroni-Holm corrected p-values indicated): loss of smell (OR: 19.06 [8.72; 41.68]; p<0.001), loss of taste (OR: 17.01 [8.49; 34.10]; p<0.001), fever (OR: 4.94 [2.87; 8.50]; p<0.001), sweats and chills (OR: 3.74 [2.31; 6.07]; p<0.001), fatigue (OR: 2.99 [1.97; 4.56]; p<0.001), cough (OR: 2.81 [1.92; 4.11]; p<0.001), muscle and joint ache (OR: 2.42 [1.46; 4.00]; p=0.005), chest tightness (OR: 2.32 [1.31; 4.11]; p=0.019), head ache (OR: 2.28 [1.46; 3.56]; p=0.003), sore throat (OR: 1.92 [1.25; 2.96]; p=0.017), and nasal congestion (OR: 1.91 [1.28; 2.85]; p=0.010). Not significant were shortness of breath, other respiratory symptoms, stomach pain, nausea and vomiting **(Table 2**). The number of symptoms reported by an individual participant served as an indicator for the intensity of the disease and was 2.18-fold higher (adjusted for sex and age, 95% CI: [1.78; 2.66]) in SARS-CoV-2-infected (IgG^+^, PCR_rep+_, PCR_new+_) compared to participants without infection (**Fig. 4A**, p<0.001). 22.22% of infected (IgG^+^, PCR_rep+_, PCR_new+_) reported no symptoms at all (**Fig. 4B**); for the other infected participants symptom numbers varied between 0 and 11 (**Fig. 4B**). IgG levels of infected study participants were not significantly associated with the number of symptoms (**Fig. 4C**).

**Table 2.**
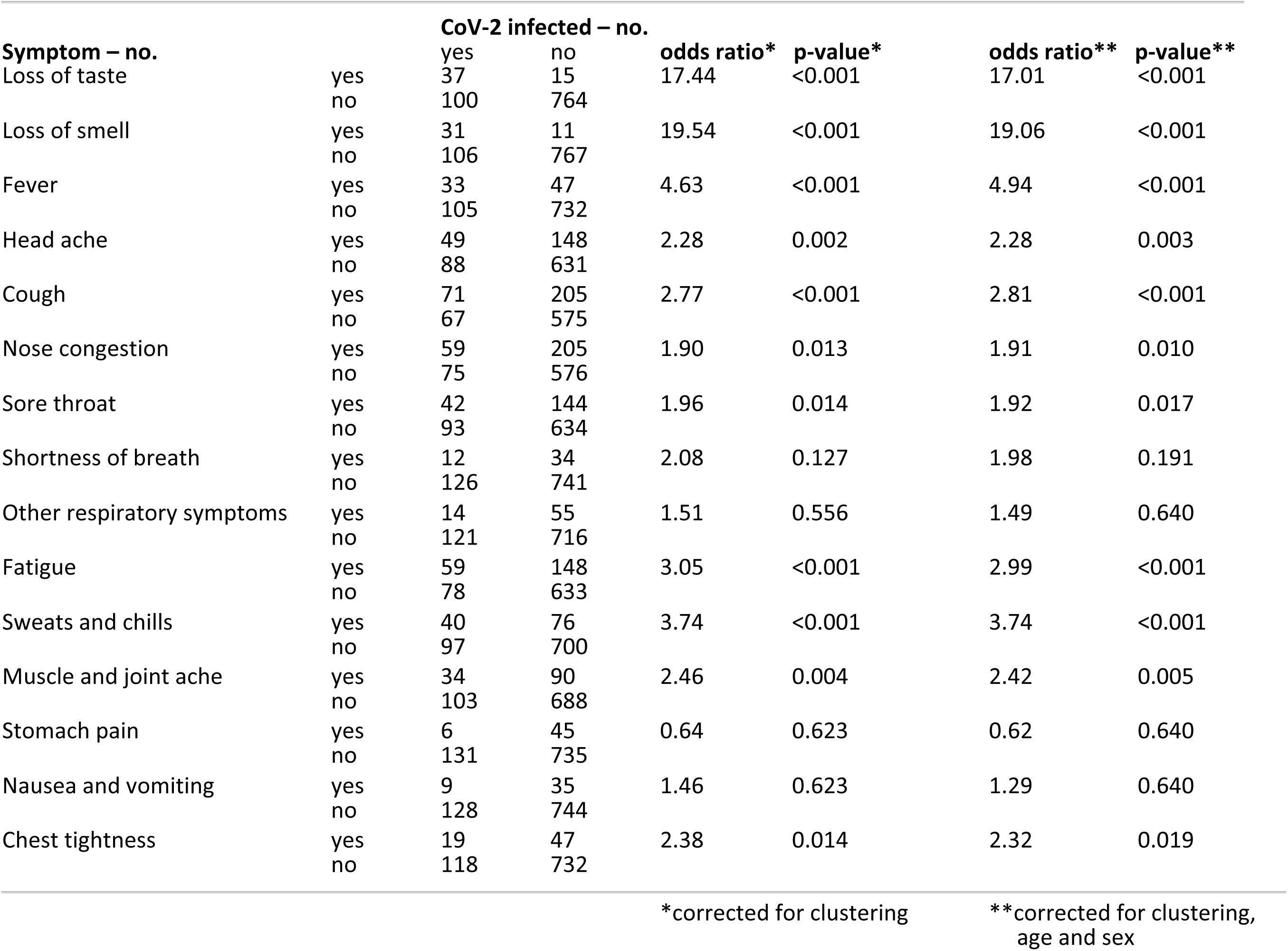
Associations between symptoms and infection rate in the 919 study participants. Missing and unknown values in the symptom variables were listwise deleted.

|  |  | CoV-2 infected – no. |  |  |  |  |  |
| --- | --- | --- | --- | --- | --- | --- | --- |
| Symptom – no. |  | yes | no | odds ratio* | p-value* | odds ratio** | p-value** |
| Loss of taste | yes | 37 | 15 | 17.44 | <0.001 | 17.01 | <0.001 |
|  | no | 100 | 764 |  |  |  |  |
| Loss of smell | yes | 31 | 11 | 19.54 | <0.001 | 19.06 | <0.001 |
|  | no | 106 | 767 |  |  |  |  |
| Fever | yes | 33 | 47 | 4.63 | <0.001 | 4.94 | <0.001 |
|  | no | 105 | 732 |  |  |  |  |
| Head ache | yes | 49 | 148 | 2.28 | 0.002 | 2.28 | 0.003 |
|  | no | 88 | 631 |  |  |  |  |
| Cough | yes | 71 | 205 | 2.77 | <0.001 | 2.81 | <0.001 |
|  | no | 67 | 575 |  |  |  |  |
| Nose congestion | yes | 59 | 205 | 1.90 | 0.013 | 1.91 | 0.010 |
|  | no | 75 | 576 |  |  |  |  |
| Sore throat | yes | 42 | 144 | 1.96 | 0.014 | 1.92 | 0.017 |
|  | no | 93 | 634 |  |  |  |  |
| Shortness of breath | yes | 12 | 34 | 2.08 | 0.127 | 1.98 | 0.191 |
|  | no | 126 | 741 |  |  |  |  |
| Other respiratory symptoms | yes | 14 | 55 | 1.51 | 0.556 | 1.49 | 0.640 |
|  | no | 121 | 716 |  |  |  |  |
| Fatigue | yes | 59 | 148 | 3.05 | <0.001 | 2.99 | <0.001 |
|  | no | 78 | 633 |  |  |  |  |
| Sweats and chills | yes | 40 | 76 | 3.74 | <0.001 | 3.74 | <0.001 |
|  | no | 97 | 700 |  |  |  |  |
| Muscle and joint ache | yes | 34 | 90 | 2.46 | 0.004 | 2.42 | 0.005 |
|  | no | 103 | 688 |  |  |  |  |
| Stomach pain | yes | 6 | 45 | 0.64 | 0.623 | 0.62 | 0.640 |
|  | no | 131 | 735 |  |  |  |  |
| Nausea and vomiting | yes | 9 | 35 | 1.46 | 0.623 | 1.29 | 0.640 |
|  | no | 128 | 744 |  |  |  |  |
| Chest tightness | yes | 19 | 47 | 2.38 | 0.014 | 2.32 | 0.019 |
|  | no | 118 | 732 |  |  |  |  |
|  |  |  |  | *corrected for clustering |  | **corrected for clustering, age and sex |  |

**Fig. 4.**
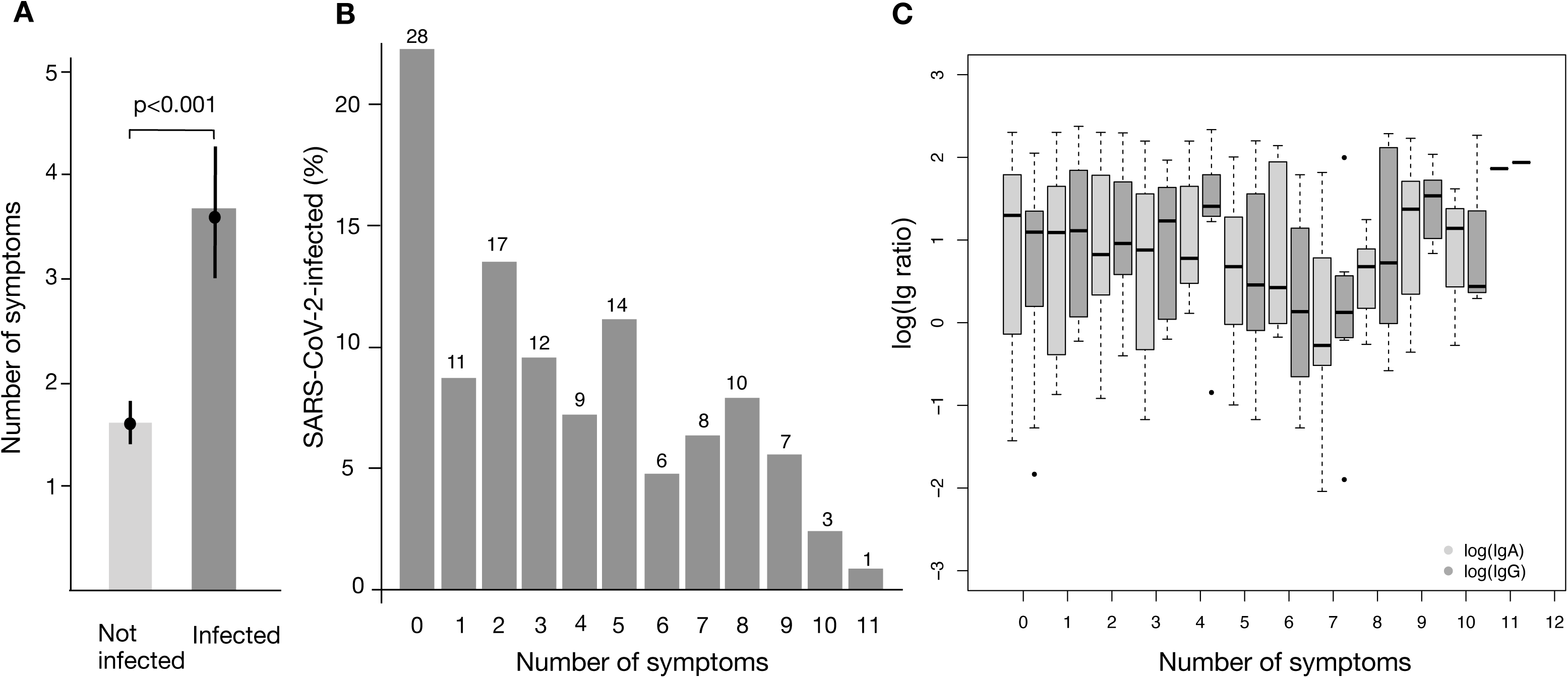
Number of symptoms and Ig in SARS-CoV-2-infected study participants. Clinical symptoms reportedly associated with SARS-CoV-2-infection were analyzed (questionnaire data). **A:** Estimated mean number of symptoms in non-infected study participants (1.61 [1.42; 1.81]) and SARS-CoV-2-infected study participants (3.58 [3.01; 4.27], Poisson GEE model, estimated relative mean increase in infected = 2.23 [1.82; 2.73], p<0.001, rho = 0.248 [0.164; 0.332]; Poisson GEE model adjusted for age and sex: estimated relative mean increase in infected = 2.18 [1.78; 2.66], p<0.001, rho = 0.250 [0.167; 0.333]). Results are based on the 876 study participants without missing values in any of the symptom items (range of the observed numbers of symptoms: 0 to 12, mean = 1.92, sd = 2.59, median = 1). Bars refer to the empirical mean values. **B:** Raw percentages of symptoms in the 126 SARS-CoV2-infected study participants without missing values for any of the symptoms. Of the SARS-CoV-2 infected, 22.22% reported that they did not have any (most left bar on x-axis: 0) of the 15 symptoms. Numbers above bars indicate the total number of individuals in the respective group. **C:** *IgA and IgG levels and intensity of symptoms*. The boxplots depict the log(IgA) (light gray) and log(IgG) (dark gray) levels in the 126 infected study participants. In a quasi-Poisson model, no significant association between the number of symptoms (response variable) and log(IgA) (covariable) was found. Similar results were obtained from a quasi-Poisson model with the number of symptoms as response variable and log(IgG) as covariable. Note: Quasi-Poisson models were used instead of Poisson GEE models because the number of households was large relative to the number of analyzed study participants (see Supplementary figure 4 A).

### Association between household size and rate of infection

SARS-CoV-2 is thought to be highly contagious. As a consequence, people living in the same household are expected to be at a much higher risk of infection. The average number of people in household clusters examined in this study was 2.27 (sd = 1.11, range 1-6) compared to Gangelt (2.44 as of 2011), the state NRW (2.02, as of Dec 2018) and Germany (1.99, as of Dec 2018). Household clusters with 5 or more people were excluded from the analysis below because of insufficient numbers (15 clusters). First, we analyzed whether the fact that an individual person was part of a one-, two-, three- or four-person household cluster changed the probability of this person being infected. We found that the infection risk was not associated with the number of people in a household cluster (**Fig. 5A**). Second, we analyzed the infection risk of a person in a household in which at least one other person was infected (**Fig. 5B**). Under the theoretical assumption that there was no increased infection risk for a second, a third or a fourth person in a household cluster in which one person was infected, the average risk in this household cluster was calculated to be 0.578 (two-person household cluster; (1 + 0.1553) / 2), 0.4369 (three-person household cluster; (1 + 2 × 0.1553) / 3) or 0.3665 (four-person household cluster; (1 + 3 × 0.1553) / 4) (**Fig. 5B**, lower gray curve). The estimated infection risk as calculated from the data was significantly above the theoretical risk without enhanced transmission (Fig. 5B, black curve, dotted lines indicate CI 95%). A significant association between household cluster size and the per-person infection risk was found (**Fig. 5B**, p<0.001). In a two-person household cluster, the estimated risk for the second infection increased from 15.53% to 43.59% [25.26%; 64.60%]; in a three-person household cluster the estimated risk for the second and third persons increased from 15.53% to 35.71% [19.57%; 55.60%] each, and in a four-person household cluster the estimated risk for the second, third and fourth persons increased from 15.53% to 18.33% [9.67%; 28.74%] each. For household clusters with at least one infected child (< 18 years), the estimated per-person risk for the other person to be infected in three-person household clusters increased from 15.53% to 66.67% [21.83%, 100.00%] and in four-person household clusters from 15.53% to 33.33% [9.02%; 71.60%].

**Fig. 5:**
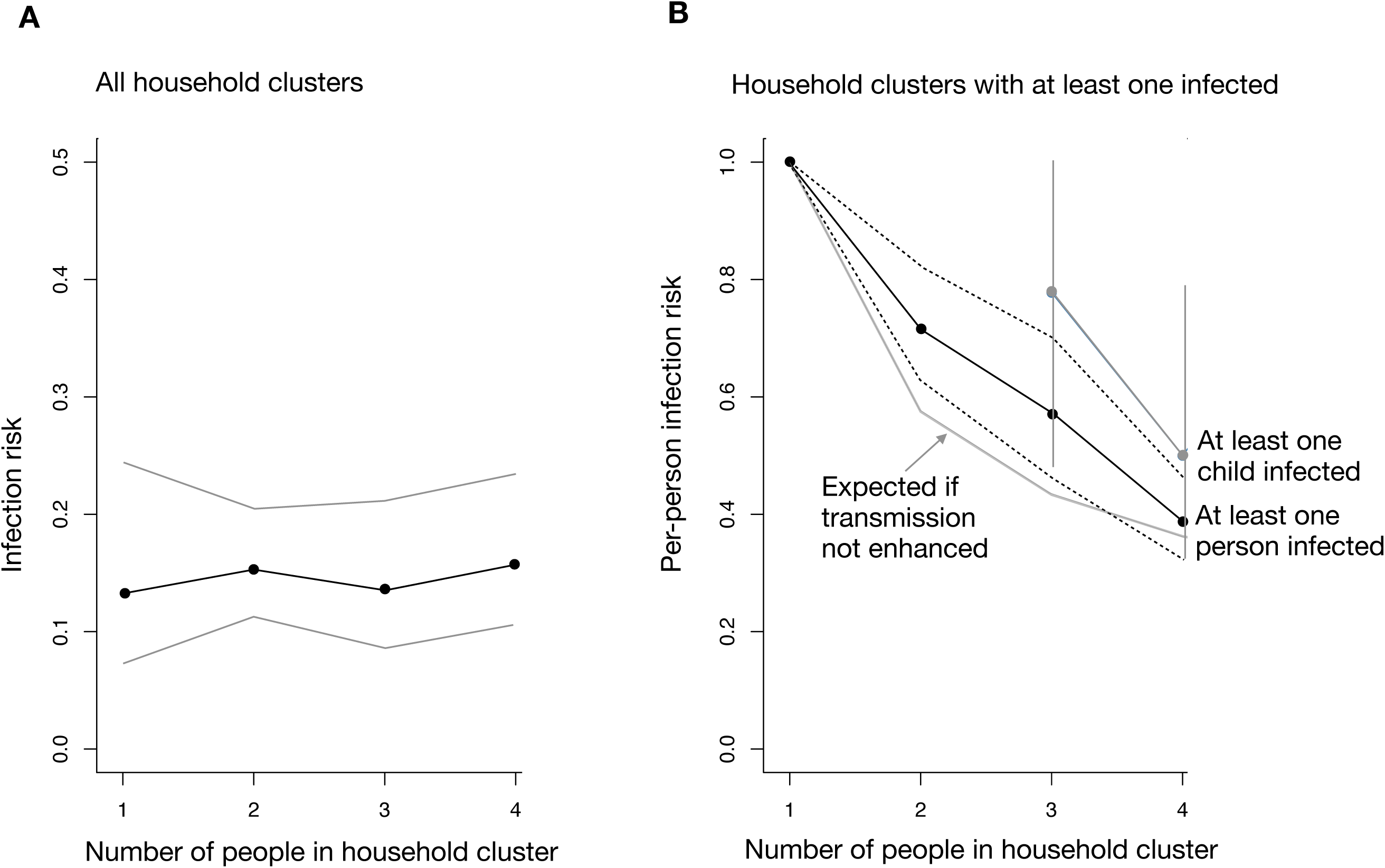
Association between household cluster size and the per-person infection risk. Due to the random selection of households, study participants were clustered within households. **A:** Estimated per-person infection risk by household cluster size (black dots; 95% CIs: gray lines). Estimates and CI limits were determined by fitting a quasi-Poisson model with the number of infected persons as response variable and household cluster size as a factor covariable. Additionally, log(household cluster size) was included as offset. No association between household cluster size and the per-person infection risk was found (p = 0.936). **B:** Per-person infection risk in household clusters in which at least one person was found infected (black curve based on 86 household clusters, with 213 persons, gray curve, based on 7 household clusters with 25 persons in which at least one infected child younger than 18 years was infected. The gray line below the black curve show the expected per-person infection risk under the assumption that there is no enhanced risk of a secondary infection in household clusters (e.g. two people in household cluster: one is infected, i.e. per-person infection risk = 1; per-person infection risk for the second person if assumed to be independent of the first person’s risk is estimated to be 0.1553 (cf. Figure 3B); expected per-person infection risk in household cluster is therefore (1 + 0.1553) / 2 = 0.578). Estimates and CI limits were determined by fitting a quasi-Poisson model with the number of infected persons as a response variable and household cluster size as a factor covariable (excluding 13 household clusters of size 1 each). Additionally, log(household cluster size) was included as an offset. A significant association between household cluster size and the per-person infection risk was found (p < 0.001). Estimates for the upper gray curve (children, CIs depicted by gray vertical lines) are based on an analogously defined model (p = 0.196). Note: 15 household clusters with more than 4 members were omitted from analysis due to small numbers. The average percentage of infected persons in these household clusters was 17.33% (0% in 9, 16.66% in 1, 20% in 2, 40% in 1, 80% in 1, 83.33% in 1 household cluster).

### Associations between sex, age, co-morbidities and super-spreading event with the rate of infection, the number of symptoms and IgA/IgG

Sex and age were not associated with the rate of infection (**Fig. 6A**). Neither IgA nor IgG of infected study participants showed significant associations with age or sex (**suppl. Fig. 3**). It is well-established that severe disease courses and fatal outcomes of SARS-CoV-2 infection are associated with the extent of underlying diseases, especially lung diseases with reduced respiratory reserves and cardiovascular diseases. We therefore analyzed the associations between co-morbidities on both the infection rate and the number of symptoms. In the questionnaire, study participants were asked to report whether they had pre-existing diseases or disease states, including lung diseases, cardiovascular diseases, neurological diseases and stroke, cancer and diabetes. Neither increased rate of infection (**Fig. 6B**) nor a higher number of symptoms were found in infected individuals (suppl. Fig. 4B). For infected study participants the self-reported use of medications queried in the questionnaire (not in figure) (ibuprofen, ACE inhibitors or AT1 agonists) all had no significant associations with the infection rate or number of symptoms. Underlying morbidities of infected study participants were not associated with Ig levels (**suppl. Fig. 5**).

**Fig. 6.**
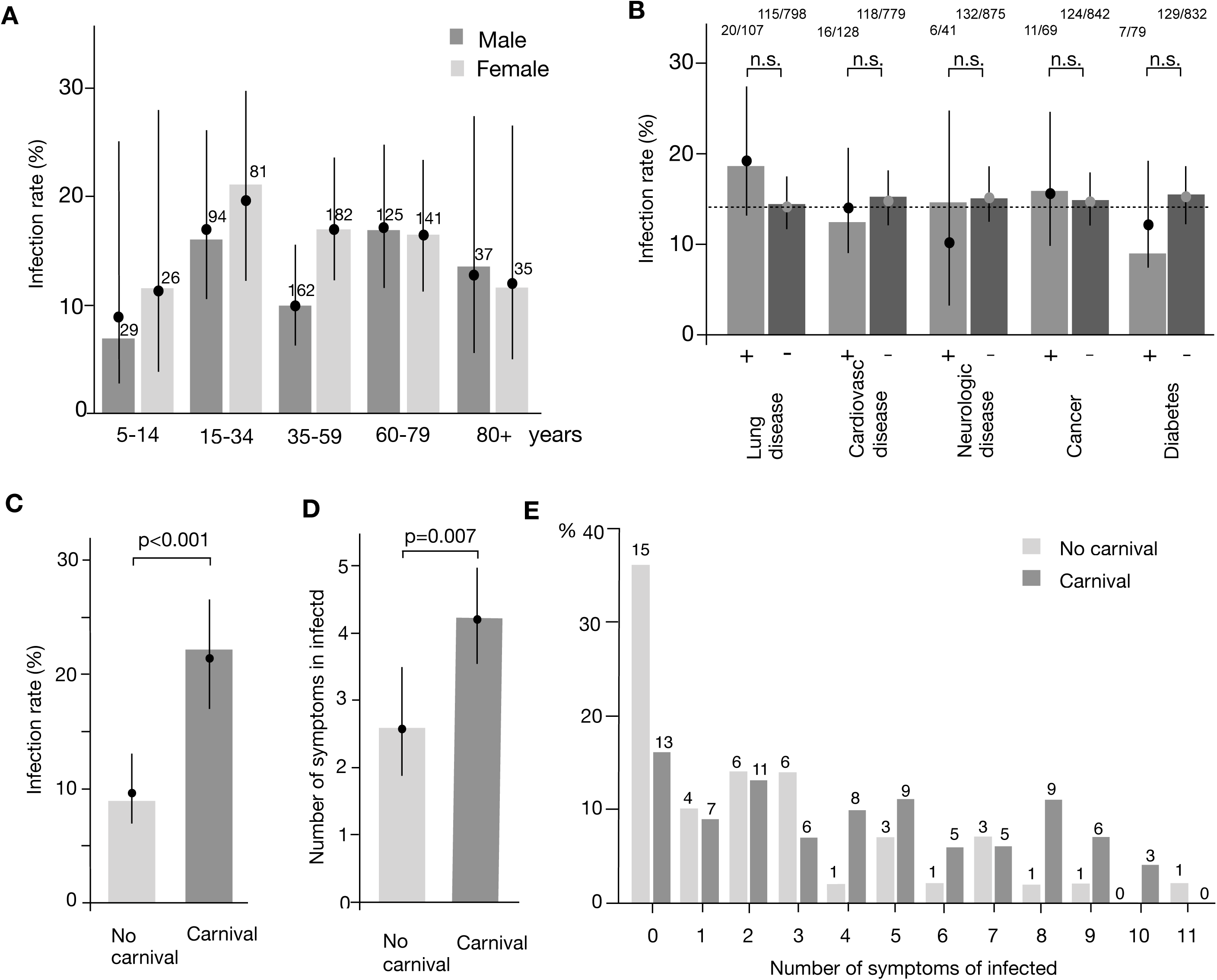
Associations of sex and age, comorbidities and super-spreading event with infection rate and symptoms. **A:** Estimated rates of infected in the study participants (filled circles, with 95% CIs) for male participants (dark gray) and female participants (light gray) stratified by age groups. Estimates were obtained by fitting logistic GEE models with the infection status as response variable and age as covariable (rho = 0.256 [0.104; 0.407] and rho = 0.244 [0.154; 0.334], respectively, in the male and female subgroups). Bars refer to the raw percentages. In a logistic GEE model with both sex and age as covariables, neither sex (OR = 1.28 [0.95; 1.73] for females, p = 0.101) nor age (OR = 1.03 [0.94; 1.14] per 10 years, p = 0.539) were found to be associated with infection status. Numbers above bars indicate the total number of individuals in the respective group. **B:** For each of the co-morbidities, the infection rate (%) was determined by fitting a logistic GEE model with infection status as response variable to the data of all study participants (light gray: co-morbidity present (+), dark gray: co-morbidity not present (-)). Point estimates obtained from the GEE models are represented by filled circles (with 95% CIs). The bars represent the raw percentages of infected in each of the subgroups (calculated from the participant numbers shown above the bars). No associations between the infection status and any of the co-morbidities were found (Bonferroni-Holm corrected p-values from GEE models at top of figure). Associations remained statistically insignificant in GEE models that included sex and age as additional covariables. Raw proportions are indicated above bars. **C:** The association with the life style factor ‘celebrating carnival’ was analyzed (questionnaire “have you celebrated carnival?” yes/no). Celebrating carnival was not limited to attending the main carnival event (Kappensitzung in Gangelt). Estimated infection rates (%; with 95% CIs) of participants not celebrating carnival (light gray) and participants celebrating carnival (dark gray). Point estimates (filled circles) and CIs were obtained by fitting a logistic GEE model with infection status as response variable and carnival (yes/no) as a factor covariable. The bars represent the raw percentage values. There was a significant positive association between celebrating carnival and infection status (OR = 2.56 [1.67; 3.93], p < 0.001, rho = 0.351 [0.162; 0.540]). Similar results were obtained when adding sex and age as covariables to the GEE model (OR = 3.08 [1.92; 4.95], p < 0.001, rho = 0.340 [0.126; 0.554]). Analyses were based on the 915 participants that had complete data in both the carnival and the infection variables. **D:** Estimated mean number of symptoms in infected participants not celebrating carnival (light gray) and in infected participants celebrating carnival (dark gray). Point estimates (filled circles) and CIs were obtained by fitting a quasi-Poisson model with the number of symptoms as response variable and carnival (yes/no) as a factor covariable. The quasi-Poisson model was used instead of a Poisson GEE model because the number of households was large relative to the number of analyzed study participants (see Supplementary figure 4 A). There was a significant positive association between celebrating carnival and the number of symptoms (estimated relative mean increase = 1.63 [1.15; 2.33], p = 0.007). Similar results were obtained when adding sex and age as covariables to the model (estimated relative mean increase = 1.62 [1.12; 2.34], p = 0.011). Analyses were based on the 124 infected participants that had complete data in both the carnival and infection variables. E: Raw percentages of infected participants celebrating carnival, grouped by their numbers of symptoms. Numbers above bars indicate the total number of individuals in the respective group.

### Associations between celebrating carnival and rate of infection and number of symptoms

The impact of super-spreading events on the dynamics of the SARS-CoV-2 pandemic is well-established^20,21^. The carnival festivities in Gangelt are mostly visited by local people staying in the area after the event, therefore providing a unique setting to study the mechanisms of super-spreading more closely than in events where people are traveling and thus disappearing from a local study population. We analyzed whether celebrating carnival (main carnival session “Kappensitzung” or other carnival festivities) was associated with the rate of infection and the intensity of the infection, based on the number of symptoms. Study participants were asked to indicate whether they had participated in carnival events. There was a significant positive association between celebrating carnival and infection (OR = 2.56 [1.67; 3.93], p < 0.001, Fig. 6C). Furthermore, there was a significant positive association between celebrating carnival and the number of symptoms in infected study participants (estimated relative mean increase: 1.63 [1.15; 2.33], p=0.007, **Fig. 6D**). While the percentage of asymptomatic infected participants was 36% without celebrating carnival, only 16% who had celebrated carnival were asymptomatic (**Fig. 6E**).

## Discussion

One key parameter to assessing the potential impact that SARS-CoV-2 infection poses on societies is the fatality rate. However, the fatality rate of ‘cases’ (case fatality rate, CFR) widely varies between countries. ‘Cases’ do not cover the whole spectrum of SARS-CoV-2 infections reaching from asymptomatic to lethal. Therefore, we set out to determine the infection fatality rate (IFR) based on the total number of SARS-CoV-2-infected individuals. We chose the German community Gangelt which had been exposed to a super-spreading event. A random population sample revealed that an estimated 15.53% of the population in this community is or was infected with the virus, which is 5-fold higher than the officially reported number of PCR-positives. Based on the estimated percentage of infected people in this population, the IFR was estimated to be 0.36% [0.29%; 0.45%]. Infection was highly associated with known characteristic symptoms of SARS-CoV-2 infection such as loss of smell and taste. The risk of being infected was not found to be associated with the number of participants living in the same household, and the estimated risk to be infected in a household cluster with one person already infected (secondary infection risk) was distinctly below 100%. The frequency of infection did not significantly differ between age groups from children to the elderly and was not found to be associated with sex. Co-morbidities such as underlying lung disease or cardiovascular disease did not show associations with the risk of infection. Notably, this does not contradict the well-established fact that co-morbidities such as lung disease predispose for severe disease outcomes^22,23^. The use of ACE-inhibiting drugs or ibuprofen did not show an association, as previously speculated^24^.

In our study, infection is defined as either PCR positive, anti-SARS-CoV2+ IgG seropositive or both, thus including present and past infections. Since SARS-CoV-2 only arrived in February, seropositives are expected to cover all infections except the very recent. This may become different as the pandemic continues, since a decrease in antibody titers over time needs to be considered in the calculation of the IFR. Furthermore, in our study, the number of reported PCR positives (2.39%) was lower than in the overall population (3.08%) of this high-prevalence community. This indicates that infected individuals may be underrepresented in our study population. Although this is plausible (no response to study request due to illness, hospital, ICU, already known infection status, etc.) and would lead to a correction by factor 1.29, we chose to use the uncorrected lower percentage to conservatively estimate the total number of infected and the resulting IFR in the population.

To determine the IFR, the collection of materials and information including the reported cases and deaths was closed at the end of the study acquisition period (April 6^th^), and the IFR was calculated based on those data. However, some of the individuals still may have been acutely infected at the end of the study acquisition period (April 6^th^) and thus may have succumbed to the infection later on. In fact, in the 2-week follow-up period (until April 20^th^) one additional COVID-19 associated death was registered. The inclusion of this additional death would bring up the IFR from 0.36% to an estimated 0.41% [0.33%; 0.52%].

Although the IFR is much less variable than the infection rate in different parts of the country, the IFR may still be affected by certain circumstances. The community in which this study was performed experienced a super-spreading event. The IFR was unlikely affected by an overwhelmed health care system because sufficient numbers of ICU beds and ventilators were available at all times. However, it is possible that the super-spreading event itself caused more severe cases. In our study, we found a highly significant increase in both infection rate and number of symptoms when people attended carnival festivities, as compared to people who did not celebrate carnival. This association with carnival was at the same level when adjusted for the age of the participants. At this point, the reason for the association with celebrating carnival remains speculative. Thus far, we could not identify confounding factors that would explain the observed difference. However, it is well established that the rate of particle emission and superemission during human speech increases with voice loudness^25^. Because of loud voices and singing in close proximity are common in carnival events, it is reasonable to speculate that a higher viral load at the time of infection caused the higher intensity of symptoms and thus more severe clinical courses of the infection. Notably, results from experimental human influenza infection studies have demonstrated that the symptom score depends on the viral dose administered^26,27^. Similar observations have been made for MERS^28^ and SARS^29^. Little is known about the infection dynamics of SARS-CoV-2. Future studies designed to specifically analyze the infection chains after super-spreading events may provide further insight. If substantiated, the IFR under strict hygiene measures might be lower than the IFR in the context of a super-spreading event in this study, with important consequences for the strategy against the pandemic. In this context, it is interesting to note that in our study, 22% of infected individuals were asymptomatic, confirming previous reports of about 20% asymptomatic carriers that contribute to the spread of infection^30-32^. Notably, asymptomatic infected individuals in our study present with substantial antibody titers. Furthermore, since the mean symptom number of non-infected in our study was 1.6 (of 15 symptoms), it would be also appropriate to count infected study participants reporting up to 1 symptom as individuals with no symptoms above the baseline level of uninfected study participants, thereby increasing the percentage of asymptomatic infected individuals to 30.1%.

Given the high contagiousness of SARS-CoV-2, one would expect high rates of transmission. However, in our study we found a relatively moderate increase of the secondary infection risk which depended on the household cluster size (increase from 15.5% baseline risk by 28% for two people, 20% for three people, 3% for four people). This finding is consistent with recent observations of secondary infection risk of 16.3% in Chinese^33^ and 7.56% in South Korea^34^. The reason for the comparably low secondary infection risk despite the high rate of transmission is currently unknown, but it is seen with other respiratory infections such as influenza (H1N1) 14.5%^35^ or SARS 14.9%^36^. Secondary household members may have acquired a level of immunity (e.g. T cell immunity) that is not detected as positive by our ELISA, but still could protect those household members from a manifest infection^26,37^.

To date, knowledge about SARS-CoV-2 immunity is rather scarce. Whether the Ig levels detected in infected individuals in our study are protective and how long such a protection lasts is not currently known. Virus neutralization control assays as performed in our study add information, but do not provide evidence for the presence of an effective immunity. As other tests, virus neutralization assays in general can be false positive, as cross-reactivity between betacoronaviruses is well-known^38,39^. Likewise a lack of virus neutralization does also not exclude a past infection as there is ample evidence that not all antibody responses neutralize but still may provide some degree of protective immunity^40,41^. Therefore, at this point our study uses IgG values as indicator whether an individual was infected but not as evidence for existing immunity. However, one may assume that a certain degree of protection might exist even if the IgG levels are below the threshold of the ELISA. Such individuals are not counted as infected in our study, yet this hidden number of infected could possibly represent an important component towards immunity in a population. The analysis of anti-SARS-CoV-2 IgM might help to further close this window in the future.

Since i) a high degree of PCR testing was performed in this community by the health authorities during the outbreak of SARS-CoV-2 infection, and ii) the outbreak was largely over, this community was chosen as an ideal site to estimate the real number of infected individuals. It is important to note that the infection rate in Gangelt is not representative for other regions in Germany or other countries. However, with the limitations discussed above, the IFR calculated here remains a useful metric for other regions with higher or lower infection rates. If in a theoretical model the here calculated IFR is applied to Germany with currently approximately 6,575 SARS-CoV-2 associated deaths (May 2nd, 2020, RKI), the estimated number of infected in Germany would be higher than 1.8 Mio (i.e. 2.2% of the German population). It will be very important to determine the true average IFR for Germany. However, because of the currently low infection rate of approximately 2% (estimated based on IFR), an ELISA with 99% specificity will not provide reliable data. Therefore, under the current non-superspreading conditions, it is more reasonable to determine the IFR in high prevalence hotspots such as Heinsberg county. The data of the study reported here will serve as baseline for follow up studies on the delta of infections and deaths to identify the corresponding IFR under those changed conditions.

## Data Availability

Data can be made available upon request

## Acknowledgement

HS, EB, MMN and GH are members of the Excellence Cluster ImmunoSensation EXC 2151 - 390873048. We would like to thank the inhabitants of Gangelt for their participation. We would also like to thank the local government of Kreis Heinsberg for their support to conduct the study. We thank Sammy Bedoui and Art Krieg for critical reading of the manuscript. Furthermore we would like to thank the following people, who helped with the study:

Gero Wilbring, Janett Wieseler, Marek Korencak, Ryan Nattrass, Jernej Pusnik, Maximilian Becker, Ann-Sophie Boucher, Marc Alexander de Boer, Rebekka Dix, Sara Dohmen, Kim Friele, Benedikt Gansen, Jannik Geier, Marie Gronemeyer, Sarah Hundertmark, Nora Jansen, Michael Jost, Louisa Khorsandian, Simon Krzycki, Ekaterina Kuskova Judith Langen, Silvia Letmathe, Ann-Kathrin Lippe, Jonathan Meinke, Freya Merker, Annika Modemann, Janine Petras, Sophie Marie Porath, Anna Quast, Laurine Reese, Isabel Maria Rehbach, Jonas Richter, Thea Rödig, Eva Schmitz, Tobias Schremmer, Louisa Sommer, Jennifer Speda, Yuhe Tang, Oliver Thanscheidt, Franz Thiele, Johanna Thiele, Julia Tholen, Sophia Tietjen, Moritz Transier, Maike van der Hoek, Tillmann Verbeek, Sophia Verspohl, Kira Vordermark, Julian Wirtz, Marina Wirtz, Lisa Zimmer, Philip Koenemann, Adi Yaser, Lisa Anna, Katharina Bartenschlager, Lisa Baum, Roxana Böhmer Romero, Diana de Braganca, Isabelle Engels, Moritz Färber, Carina Fernandez Gonzalez, Lucia Maria Goßner, Victoria Handschuch, Franziska Georgia Liermann, Steffan Meißner, Laura Racenski, Patrick, Denis Raguse, Larissa Reiß, Maximilian Rölle, Franziska Scheele, Chiara Schwippert, Arlene Christin Schwippert, Antonia Seifert, Joshua David Stockhausen, Sofia Waldorf, Leonie Weinhold, Nicolai Trimpop, Julia Reinhardt, Vera Gast, Michelle Yong, Eva Engels, Jonathan Meinke, Susanne Schmidt, Janine Schulte, Saskia Schmitz, Kübra Bayrak, Regina Frizler, Katarzyna Andryka, Soía Soler, Thomas Zillinger, Marcel Renn, Patrick Müller, Dillon Corvino, Zeinab Abdullah, Katrin Paeschke, Hiroki Kato, Daniel Hinze, Martina Schmidt, Arcangelo Ricchiuto, Sonja Gross, Uta Wolber, Marion Zerlett, Esther Sib, Benjamin Marx, Souhaib Aldabbagh. We thank Stefan Holdenrieder, Alexander Semaan, Bernd Pötzsch and Georg Nickening for providing control samples.

---

Supplementary figures

**Supplementary figure 1:**
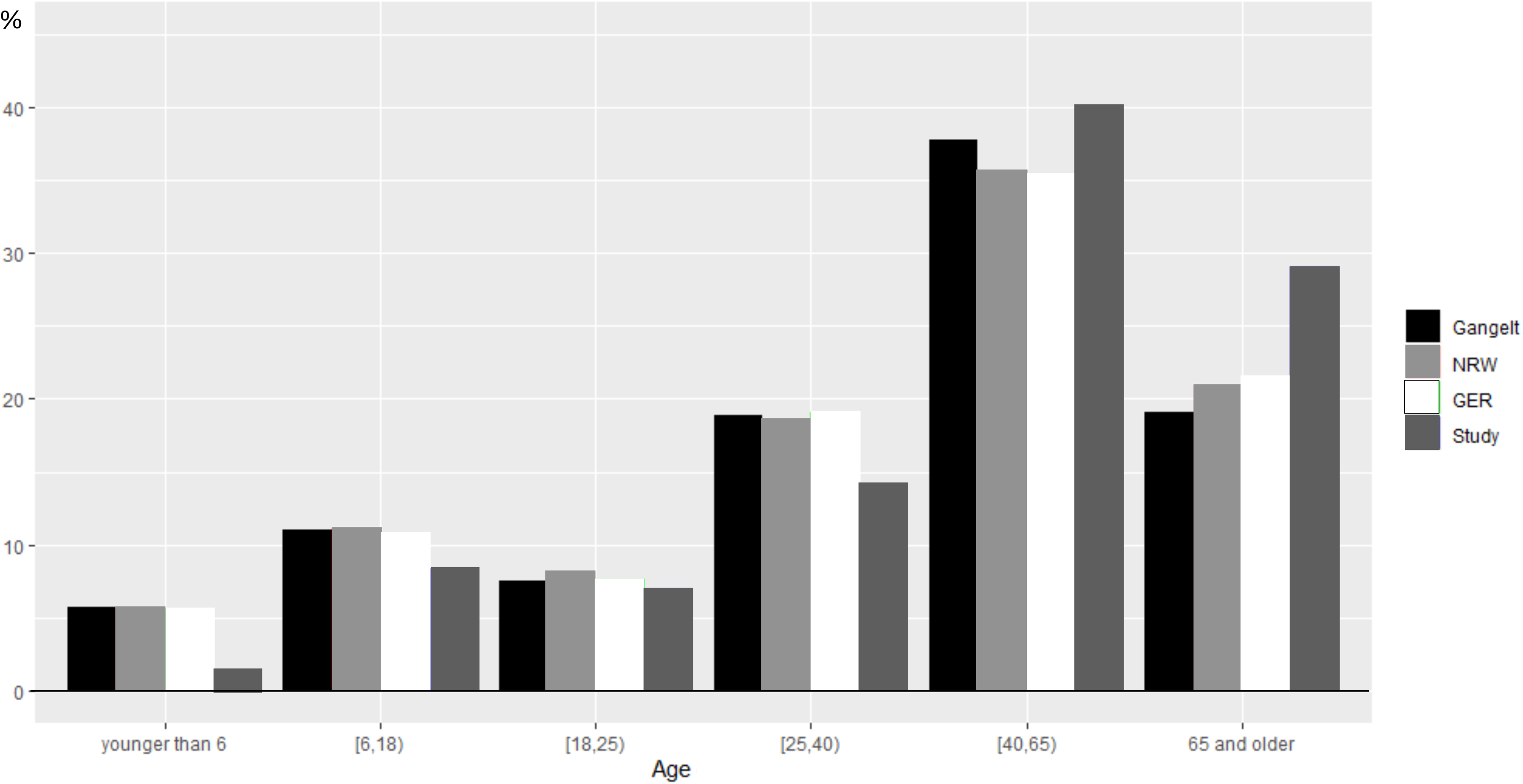
Comparison of the distribution of age groups of the study participants to those in the community of Gangelt, the state NRW and Germany. Data were obtained from the Landesdatenbank NRW (reporting date: December 31, 2017) and statista.com (reporting date: December 31, 2018).

**Supplementary figure 2:**
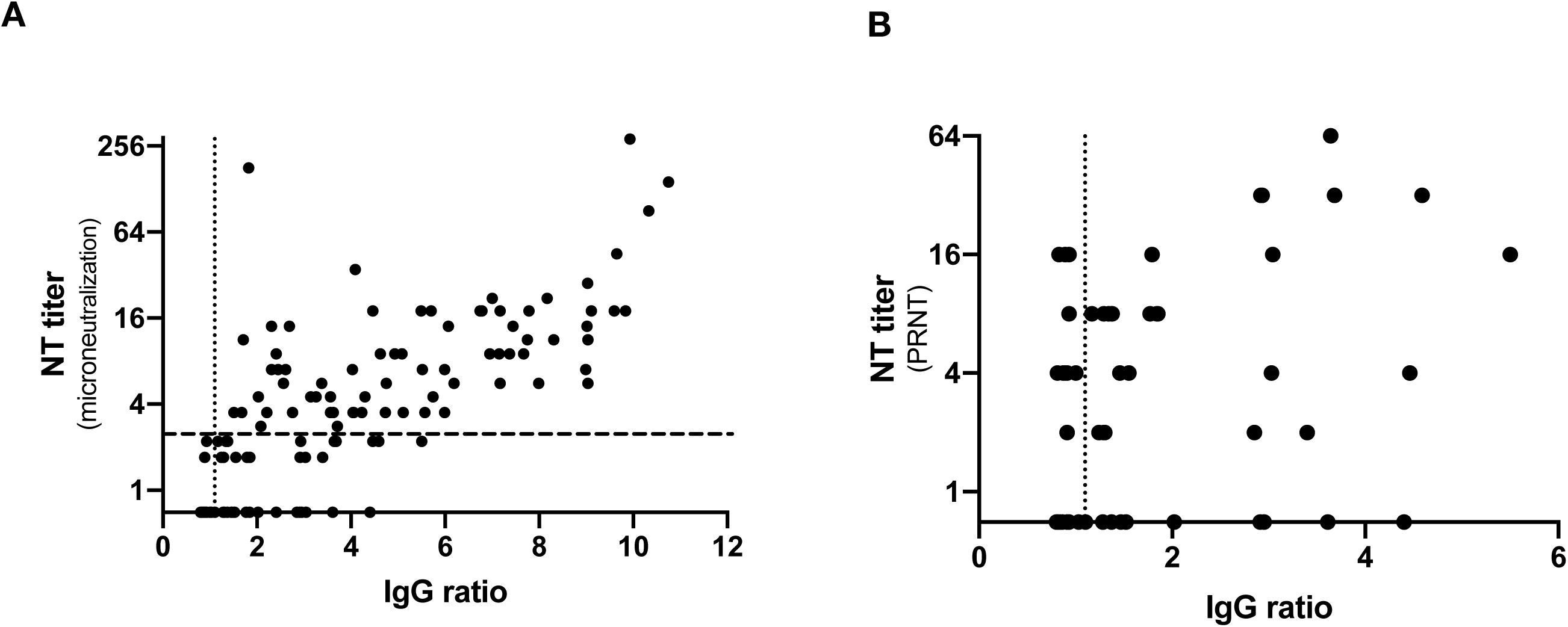
Comparison of IgG levels to neutralization activity. **A:** NT titers were determined by a microneutralization assay using 100 TCID_50_. Titers indicate the reciprocal value of the plasma dilutions that protect 50% of the wells at incubation with 100 TCID_50_. Samples able to suppress the cytopathic effect (CPE) in at least all three wells of the 1:2 dilution (NT titer ≥ 2.8) are depicted above the dashed line. Samples for which the CPE was suppressed in one or two wells of the 1:2 dilution are shown directly below the dashed line. Samples showing a CPE in all wells with either equal or reduced severity compared to the negative control were depicted at the level of the x-axis. **B:** Samples below the dashed line in A were re-evaluated using a plaque reduction neutralization test (PRNT). The neutralizing titers were calculated as the reciprocal of serum dilutions resulting in neutralization of 50% input virus (NT_50_). Samples without neutralizing activity in the NT_50_ assay were depicted at the level of the x-axis. Dotted line: upper borderline for ELISA IgG ratio.

**Supplementary figure 3:**
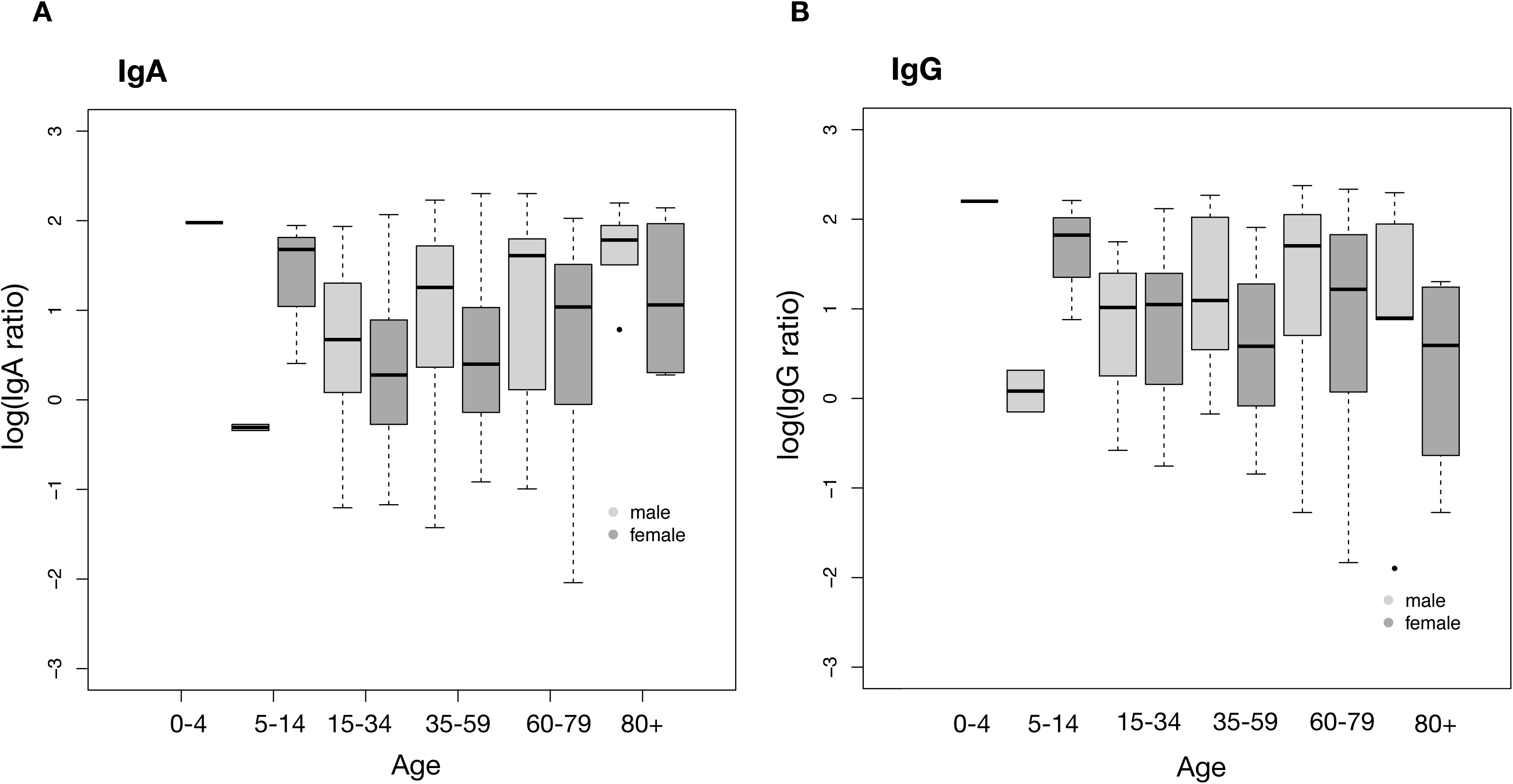
IgA and IgG levels in study participants of different sex and age. The boxplots depict the log(IgA) **(A**) and log(IgG) (**B**) levels of the infected study participants grouped by sex and age. No significant associations were found (i) between log(IgA), log(IgG), and sex in Gaussian models with log(IgA) and log(IgG) as response variables and sex as covariable, and (ii) between log(IgA), log(IgG), sex, age in Gaussian models with log(IgA) and log(IgG) as response variables and both sex and age as covariables. Note: Gaussian models were used instead of GEE models because the number of household clusters was large relative to the number of analyzed study participants, see Supplementary figure 4 A.

**Supplementary figure 4:**
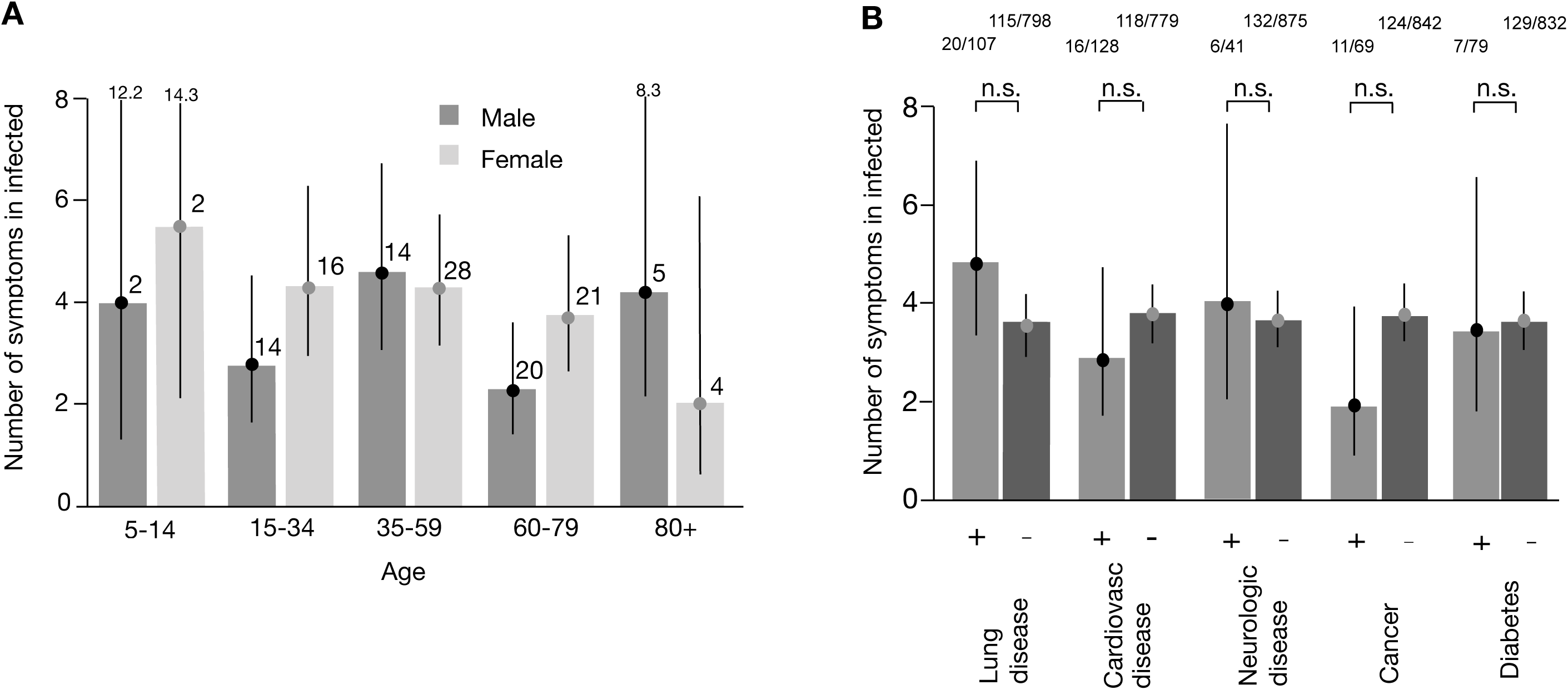
Associations of sex and age and co-morbidities with disease intensity. **A:** Estimated mean numbers of symptoms (filled circles, with 95% CIs) in 55 infected male participants (living in 52 household clusters, dark gray) and 71 infected female participants (living in 66 household clusters, light gray) stratified by age groups. In view of the large number of household clusters relative to the number of persons (52/55, 66/71), estimates were obtained by fitting non-GEE quasi-Poisson models with the number of symptoms as response variable and age as covariable in the male and female subgroups. Bars refer to the empirical mean values. In a quasi-Poisson model with both sex and age as covariables, neither sex (estimated relative mean increase = 1.26 [0.93; 1.71] for females, p = 0.142) nor age (estimated relative mean increase = 0.97 [0.90; 1.04] per 10 years, p = 0.348) were found to be associated with the number of symptoms. Results are based on the 126 infected study participants without missing values in any of the symptom items. Numbers above bars indicate the total number of individuals in the respective group. Note: There were no children aged less than 5 years. Numbers above CIs indicate the upper CI limits. **B:**r each of the co-morbidities, the mean number of symptoms in the infected participants was determined by fitting a non-GEE quasi-Poisson model with the number of symptoms as response variable to the data of the infected study participants (light gray: co-morbidity present (+), dark gray: co-morbidity not present (-)). Quasi-Poisson models were used instead of GEE models for the same reason as in A. Results are based on the 126 infected study participants without missing values in any of the symptom items. Point estimates obtained from the models are represented by filled circles (with 95% CIs). The bars represent the empirical means of the number of symptoms in each of the subgroups. Raw proportions numbers in each of the subgroups are shown above the bars. Numbers above error bars indicate the upper end of the error bar. No significant associations between the presence of any of the co-morbidities and the number of symptoms were found (Bonferroni-Holm corrected). Analogous results were found when sex and age were added as covariables to the quasi-Poisson models.

**Supplementary figure 5:**
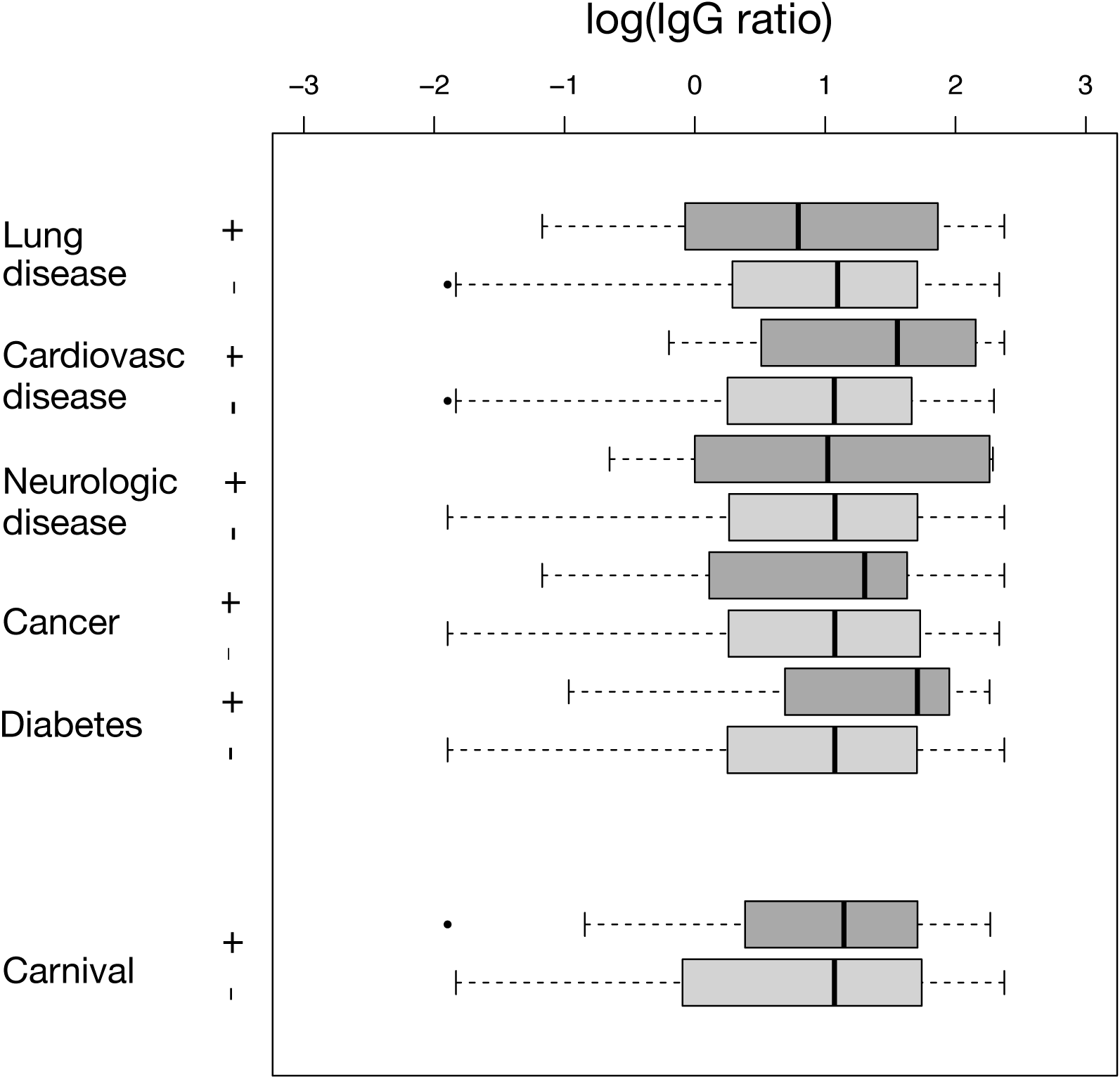
Associations between IgG levels, co-morbidities, and celebrating carnival in the infected study participants. The boxplots depict the log(IgG) levels of the infected study participants in subgroups defined by the presence of co-morbidities and celebrating carnival. In Gaussian models with log(IgG) as response variable (used instead of GEE models for the same reason as in Supplementary figure 4 A), no associations between log(IgG), the co-morbidities, and celebrating carnival were found.

**Supplementary Table 1.** Characteristics of the 88 study participants not evaluable for infection status.

| Characteristic | Value |
| --- | --- |
| Participants (no., %) living in household clusters of size |  |
| 2 | 11 (12.50) |
| 3 | 29 (32.95) |
| 4 | 28 (31.82) |
| ≥ 5 | 20 (22.73) |
| Age |  |
| Median (range) | 9 yr (1 yr – 84 yr) |
| Distribution – no. (%) |  |
| <5 yr | 28 (31.82) |
| 5-14 yr | 28 (31.82) |
| 15-34 yr | 12 (13.64) |
| 35-59 yr | 10 (11.36) |
| 60-79 yr | 9 (10.23) |
| >79 yr | 1 (1.14) |
| Sex – no. (%) |  |
| Male | 36 (41.38) |
| Female | 50 (57.47) |
| Diverse | 1 (1.15) |
| Missings: 1 |  |
| PCR<sub>reported</sub> – no. (%) |  |
| Yes | 13 (14.78) |
| No | 75 (85.22) |
| PCR<sub>reported</sub> positive – no (%) |  |
| Yes | 0 (0.00) |
| No | 13 (100.00) |
| Carnival – no (%) |  |
| Yes | 46 (52.27) |
| No | 42 (47.73) |

## References

1. Huang C, Wang Y, Li X, et al. Clinical features of patients infected with 2019 novel coronavirus in Wuhan, China. Lancet 2020;395:497–506.

2. Chen N, Zhou M, Dong X, et al. Epidemiological and clinical characteristics of 99 cases of 2019 novel coronavirus pneumonia in Wuhan, China: a descriptive study. Lancet 2020;395:507–13.

3. Coronavirus (COVID-19) Dashboard. 2020. (Accessed at https://covid19.who.int.)

4. Shujuan Ma, Jiayue Zhang, Minyan Zeng, et al. Epidemiological parameters of coronavirus disease 2019: a pooled analysis of publicly reported individual data of 1155 cases from seven countries. medRxiv 2020032120040329; doi: https://doiorg/101101/2020032120040329 2020.

5. Lavezzo E, Franchin E, Ciavarella C, et al. Suppression of COVID-19 outbreak in the municipality of Vo, Italy. 2020.

6. Bi Q, Wu Y, Mei S, et al. Epidemiology and Transmission of COVID-19 in Shenzhen China: Analysis of 391 cases and 1,286 of their close contacts. 2020.

7. Statistics and Research Coronavirus Disease (COVID-19). 2020. (Accessed at (https://ourworldindata.org/coronavirus#what-do-we-know-about-the-risk-of-dying-from-covid-19)..)

8. Yongchen Z, Shen H, Wang X, et al. Different longitudinal patterns of nucleic acid and serology testing results based on disease severity of COVID-19 patients. Emerg Microbes Infect 2020: 1–14.

9. Wolfel R, Corman VM, Guggemos W, et al. Virological assessment of hospitalized patients with COVID-2019. Nature 2020.

10. Okba NMA, Muller MA, Li W, et al. SARS-CoV-2 specific antibody responses in COVID-19 patients. 2020.

11. Corman VM, Landt O, Kaiser M, et al. Detection of 2019 novel coronavirus (2019-nCoV) by real-time RT-PCR. Euro Surveill 2020;25.

12. Ge XY, Li JL, Yang XL, et al. Isolation and characterization of a bat SARS-like coronavirus that uses the ACE2 receptor. Nature 2013;503:535–8.

13. Ramakrishnan MA. Determination of 50% endpoint titer using a simple formula. World J Virol 2016;5:85–6.

14. Population-based age-stratified seroepidemiological investigation protocol for COVID-19 virus infection. 2020. (Accessed at https://www.who.int/publications-detail/population-based-age-stratified-seroepidemiological-investigation-protocol-for-covid-19-virus-infection.)

15. Hanley JA, Negassa A, Edwardes MD, Forrester JE. Statistical analysis of correlated data using generalized estimating equations: an orientation. Am J Epidemiol 2003;157:364–75.

16. Rogan WJ, Gladen B. Estimating prevalence from the results of a screening test. Am J Epidemiol 1978;107:71–6.

17. C. DA, V. HD. Bootstrap methods and their application.: Cambridge University Press; 1978.

18. Global Covid-19 Case Fatality Rates. 2020. (Accessed at https://www.cebm.net/covid-19/global-covid-19-case-fatality-rates.)

19. Rothman KJ, Greenland S, Lash TL. Modern Epidemiology. 3 ed: Wolters Kluwer; 2008.

20. Liu Y, Eggo RM, Kucharski AJ. Secondary attack rate and superspreading events for SARS-CoV-2. Lancet 2020;395:e47.

21. Al-Tawfiq JA, Rodriguez-Morales AJ. Super-spreading events and contribution to transmission of MERS, SARS, and COVID-19. J Hosp Infect 2020.

22. Feng Y, Ling Y, Bai T, et al. COVID-19 with Different Severity: A Multi-center Study of Clinical Features. Am J Respir Crit Care Med 2020.

23. Wang T, Du Z, Zhu F, et al. Comorbidities and multi-organ injuries in the treatment of COVID-19. Lancet 2020;395:e52.

24. Fang L, Karakiulakis G, Roth M. Are patients with hypertension and diabetes mellitus at increased risk for COVID-19 infection? Lancet Respir Med 2020;8:e21.

25. Asadi S, Wexler AS, Cappa CD, Barreda S, Bouvier NM, Ristenpart WD. Aerosol emission and superemission during human speech increase with voice loudness. Sci Rep 2019;9:2348.

26. Wilkinson TM, Li CK, Chui CS, et al. Preexisting influenza-specific CD4+ T cells correlate with disease protection against influenza challenge in humans. Nat Med 2012;18:274–80.

27. Memoli MJ, Czajkowski L, Reed S, et al. Validation of the wild-type influenza A human challenge model H1N1pdMIST: an A(H1N1)pdm09 dose-finding investigational new drug study. Clin Infect Dis 2015;60:693–702.

28. Oh MD, Park WB, Choe PG, et al. Viral Load Kinetics of MERS Coronavirus Infection. N Engl J Med 2016;375:1303–5.

29. Hung IF, Cheng VC, Wu AK, et al. Viral loads in clinical specimens and SARS manifestations. Emerg Infect Dis 2004;10:1550–7.

30. Lai CC, Liu YH, Wang CY, et al. Asymptomatic carrier state, acute respiratory disease, and pneumonia due to severe acute respiratory syndrome coronavirus 2 (SARS-CoV-2): Facts and myths. J Microbiol Immunol Infect 2020.

31. Mizumoto K, Kagaya K, Zarebski A, Chowell G. Estimating the asymptomatic proportion of coronavirus disease 2019 (COVID-19) cases on board the Diamond Princess cruise ship, Yokohama, Japan, 2020. Euro Surveill 2020;25.

32. Bai Y, Yao L, Wei T, et al. Presumed Asymptomatic Carrier Transmission of COVID-19. JAMA 2020.

33. Li W, Zhang B, Lu J, et al. The characteristics of household transmission of COVID-19. Clin Infect Dis 2020.

34. Covid-19 National Emergency Response Center E, Case Management Team KCfDC, Prevention. Coronavirus Disease-19: Summary of 2,370 Contact Investigations of the First 30 Cases in the Republic of Korea. Osong Public Health Res Perspect 2020;11:81–4.

35. Carcione D, Giele CM, Goggin LS, et al. Secondary attack rate of pandemic influenza A(H1N1) 2009 in Western Australian households, 29 May-7 August 2009. Euro Surveill 2011;16.

36. Lau JT, Lau M, Kim JH, Tsui HY, Tsang T, Wong TW. Probable secondary infections in households of SARS patients in Hong Kong. Emerg Infect Dis 2004;10:235–43.

37. Braun J, Loyal L, Frentsch M, et al. 2020.

38. Meyer B, Drosten C, Muller MA. Serological assays for emerging coronaviruses: challenges and pitfalls. Virus Res 2014;194:175–83.

39. Chan JF, Lau SK, To KK, Cheng VC, Woo PC, Yuen KY. Middle East respiratory syndrome coronavirus: another zoonotic betacoronavirus causing SARS-like disease. Clin Microbiol Rev 2015;28:465–522.

40. Ilinykh PA, Huang K, Santos RI, et al. Non-neutralizing Antibodies from a Marburg Infection Survivor Mediate Protection by Fc-Effector Functions and by Enhancing Efficacy of Other Antibodies. Cell Host Microbe 2020.

41. Corey L, Gilbert PB, Tomaras GD, Haynes BF, Pantaleo G, Fauci AS. Immune correlates of vaccine protection against HIV-1 acquisition. Sci Transl Med 2015;7:310rv7.

